# Prediction of confirmed, hospitalized, and severe COVID-19 cases and mechanistic insights from viral concentrations and variant dynamics in wastewater

**DOI:** 10.64898/2026.03.18.26348767

**Authors:** Michio Murakami, Ryo Watanabe, Ryo Iwamoto, Ung-il Chung, Masaaki Kitajima, Byung-Kwang Yoo

**Affiliations:** Center for Infectious Disease Education and Research (CiDER), The University of Osaka, Suita, Osaka, Japan; EIPM Center, The University of Osaka, Suita, Osaka, Japan; Graduate School of Health Innovation, Kanagawa University of Human Services, Kawasaki, Japan; AdvanSentinel Inc., 7-6 Doshomachi 4-Chome, Chuo-ku, Osaka, Osaka, Japan; Department of Bioengineering, Graduate School of Engineering, The University of Tokyo, Tokyo, Japan; Research Center for Water Environment Technology, School of Engineering, The University of Tokyo, Bunkyo-ku, Tokyo, Japan; Faculty of Human Sciences, School of Human Sciences, Waseda University, Tokorozawa City, Saitama, Japan

**Keywords:** COVID-19, hospitalization, SARS-CoV-2, variant, wastewater-based epidemiological monitoring

## Abstract

**Background:** Following the end of a public health emergency of international concern, divergence emerged between reported coronavirus disease 2019 (COVID-19) cases and severe acute respiratory syndrome coronavirus 2 (SARS-CoV-2) RNA concentrations in wastewater. Exploring viral, clinical, patient, and surveillance-related factors underlying this divergence, we developed models to predict clinically confirmed infections, hospitalizations, and severe cases.

**Methods:** In this observational study, we analyzed ∼2 years of data from January 2022 in Kanagawa Prefecture, Japan, assessing associations between wastewater SARS-CoV-2 RNA concentrations and confirmed, hospitalized, and severe cases, adjusting for wave and variant effects.

**Findings:** Our models based on wastewater viral RNA concentrations showed high predictive accuracy (*R²* = 0.8199–0.9961), closely tracking confirmed, hospitalized, and severe cases. Models derived from earlier waves were applied to subsequent waves with residual correction based on prior prediction errors and maintained good predictive performance (root mean square error = 0.0665–0.2065). Divergence between wastewater viral RNA concentrations and reported cases was not explained by changes in viral shedding. Declines in patients’ healthcare-seeking behavior and testing were associated with trends in confirmed cases, whereas milder clinical presentation was associated with severe case trends. The lineages XBB.1.9.2 and BA.2.86 were identified as candidates associated with reduced virulence.

**Interpretation:** By incorporating understanding of viral, clinical, and surveillance-related mechanisms, wastewater surveillance may enable prediction of case trends approximately one week earlier than official reporting and inform healthcare capacity planning.

## Introduction

Coronavirus disease 2019 (COVID-19) remains a significant public health challenge even after the World Health Organization declared the end of the public health emergency of international concern (end-PHEIC) on May 5, 2023. Before the end-PHEIC declaration, the cumulative number of confirmed COVID-19 cases had reached approximately 765 million, with 6.93 million deaths ^1^. Although confirmed cases declined after the end-PHEIC declaration, countries that maintain official death statistics continue to report substantial death tolls (e.g., 48,000 in the United States and 36,000 in Japan in 2024) ^2,3^. Predicting surges in confirmed cases (i.e., infection confirmed by clinical tests), hospitalized cases, and severe cases remains essential to ensure adequate healthcare capacity ^4^.

Wastewater surveillance enables early detection of community-level COVID-19 infections by measuring severe acute respiratory syndrome coronavirus 2 (SARS-CoV-2) RNA in untreated wastewater at treatment plants ^5,6^ and has also been used to estimate hospitalized and severe cases ^7,8^. Particularly after the end-PHEIC declaration, when delays in case reporting became more common, the importance of wastewater surveillance as an early indicator of confirmed, hospitalized, and severe cases has increased.

Notably, after the end-PHEIC declaration, reported case counts have declined in relation to wastewater SARS-CoV-2 RNA concentrations ^9^. In Japan, this divergence became especially pronounced following the reclassification of COVID-19 under the Infectious Diseases Control Law on May 8, 2023, from notifiable disease surveillance (universal all-case reporting) to sentinel surveillance (partial, representative reporting) ^10^. Adjustment for hospital testing rates substantially attenuates this discrepancy ^11^, suggesting that the divergence reflects structural changes in case ascertainment rather than epidemiological dynamics alone. Potential contributors include (1) altered viral load shedding associated with lineage replacement ^12^; (2) reduced healthcare-seeking behavior for mild illness ^13^; and (3) decreased testing intensity in healthcare settings ^14^; (4) attenuation of disease severity due to viral evolution (i.e., reduced virulence) or immunity from prior infection or vaccination ^15^.

These mechanisms are likely to vary across epidemic waves and SARS-CoV-2 lineages, indicating that case-to-wastewater ratios reflect a complex interplay of viral, clinical, patient, and surveillance-related factors. Importantly, while fluctuations in viral shedding are expected to influence all case-to-wastewater ratios similarly, changes in symptom severity disproportionately affect the severe-case ratio, whereas reductions in healthcare-seeking and testing primarily influence the confirmed-case ratio. Wastewater analysis also enables estimation of variant proportions. Considering wave- and lineage-specific dynamics provides a framework to disentangle true epidemiological changes from surveillance artifacts. To our knowledge, no such integrated analyses have been conducted to date.

Therefore, our primary aim was to evaluate whether confirmed, hospitalized, and severe COVID-19 cases could be predicted based on wastewater viral RNA concentrations over an approximately two-year period encompassing five waves, from Week 1 of 2022, when the Omicron variant emerged, to Week 13 of 2024. Furthermore, by examining wave- and lineage-specific case-to-wastewater relationships, our secondary aim was to clarify the mechanisms underlying observed discrepancies between case counts and wastewater SARS-CoV-2 RNA concentrations and to establish a framework for interpreting wastewater surveillance in the post-PHEIC era.

## Methods

### Ethics

This study was approved by the Ethics Committee of Kanagawa University of Human Services (SHI No. 55; https://www.kuhs.ac.jp/research/ethics_kawasaki/).

### COVID-19 confirmed, hospitalized, and severe cases

In this observational study, data on confirmed, hospitalized, and severe COVID-19 cases in Kanagawa Prefecture, Japan were used.

During the notifiable disease surveillance period (hereafter referred to as the pre-reclassification period), the total number of newly confirmed cases was obtained ^16^. During the sentinel surveillance period (hereafter referred to as the post-reclassification period), the number of newly confirmed cases per sentinel medical institution was obtained ^17^. Data from the pre-reclassification period were converted to the number of cases per epidemiological week.

For hospitalized and severe cases, prevalence data were available ^16^. Here, hospitalized cases do not include those recovering at home or in lodging facilities. To align with the reporting schedule of the post-reclassification data, Tuesday values were used for the pre-reclassification period. Because data for May 9, 2023 were unavailable, the most recent available value (May 7, 2023) was used instead.

SARS-CoV-2 viral shedding in stools of infected individuals declines substantially after infection ^6,18^. In addition, wastewater SARS-CoV-2 RNA concentrations have been reported to correlate more strongly with incidence than with prevalence at the community level ^19^. Therefore, considering the relationship with length of hospital stay, prevalence was converted to incidence using equations (1) and (2) ^20^.

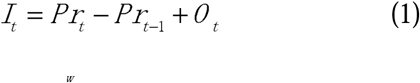

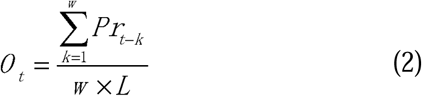

where *t* denotes time, *I* denotes weekly incidence (newly hospitalized or severe cases), *Pr* denotes weekly prevalence (number of hospitalized or severe cases), *O* denotes the weekly average outflow from the hospitalized population, *L* denotes the mean length of hospital stay (in weeks), and *w* denotes the number of weeks used in the average (*w* = *L*).

A nationwide large-scale survey reported that the median length of hospital stay for COVID-19 in Japan was 25 days during the pre-reclassification period and 19 days during the post-reclassification period ^21^. Accordingly, *L* was set to 4 and 3 weeks for the pre- and post-reclassification periods, respectively. Severe cases generally involve approximately one week of intensive care unit stay ^22,23^, resulting in longer total hospitalization periods. Therefore, for severe cases, *L* was set to 5 weeks for the pre-reclassification period and 4 weeks for the post-reclassification period. However, because some studies have reported shorter hospital stays (10–15 days) for COVID-19 patients in Japan ^22,23^, sensitivity analyses were also conducted using *L* = 1–5 weeks for hospitalized cases and *L* = 2–6 weeks for severe cases.

### Wastewater sampling and analysis

Wastewater samples were collected by grab sampling at 9:30 AM from two wastewater treatment plants in Kanagawa Prefecture. The combined catchment population was approximately 1.80 million (about 20% of the prefectural population), and the sewer systems included both separate and combined systems. In general, two samples were collected per plant per week (four samples per week in total).

Samples collected from Week 50 of 2021 (December 13, 2021) to Week 13 of 2024 were analyzed (n = 448). SARS-CoV-2 RNA and pepper mild mottle virus (PMMoV) RNA were quantified using the EPISENS-S method ^24^. Briefly, RNA was extracted from centrifuged solids, followed by one-step RT-preamplification and qPCR targeting the CDC N1 region. Viral copy numbers were determined using plasmid-based standard curves with appropriate controls. The theoretical limit of detection was 93 copies/L.

Variants were generally measured in two samples per week beginning in Week 3 of 2022. Variant proportions were determined using a targeted amplicon sequencing workflow with nested PCR targeting the spike receptor-binding domain (RBD), as previously described by Kuroiwa et al. ^25^. Briefly, indexed amplicon libraries were sequenced on an Illumina MiSeq platform. Paired-end reads were primer-trimmed, quality-filtered, denoised, and merged, then aligned to the reference spike sequence, translated, and assigned to variants based on predefined combinations of amino acid substitutions to estimate variant frequencies in wastewater samples.

### Statistical analysis

We first assessed whether wastewater SARS-CoV-2 RNA concentrations, SARS-CoV-2 RNA/PMMoV RNA ratios, and the incidence of confirmed, hospitalized, and severe cases followed normal or log-normal distributions. Among the 448 measurements of SARS-CoV-2 RNA in wastewater, 51 were below the LOD. One week of the incidence of hospitalized cases and 24 weeks of the incidence of severe cases had negative values. Considering the LOD for wastewater SARS-CoV-2 RNA as 93 copies/L and that the minimum reported value (MRV) for the incidence of hospitalized and severe cases was less than one person per week, distribution estimation was performed with left-censoring before and after log-transformation using the R package NADA ^26,27^, and the values corresponding to one-half of the non-detect proportion were estimated ^10^. Before log-transformation, the estimated values corresponding to one-half of the non-detect proportion were negative. After log-transformation, the estimated values were 129 copies/L for SARS-CoV-2 RNA in wastewater, 11.93 persons/week for the incidence of hospitalized cases, and 0.80 persons/week for the incidence of severe cases (Table S1). Accordingly, for non-log-transformed data, values below LOD or MRV were set to 0. For log-transformed data, values below LOD or MRV were set to LOD (93 copies/L) for wastewater SARS-CoV-2 RNA, MRV (1 person/week) for the incidence of hospitalized cases, and 0.80 persons/week for the incidence of severe cases.

After these replacements, Q-Q plots were used to examine whether wastewater SARS-CoV-2 RNA concentration, SARS-CoV-2 RNA/PMMoV RNA ratio, and the incidence of confirmed, hospitalized, and severe cases were closer to normal or log-normal distributions (Figure S1), as the sample size was sufficiently large ^28^. All variables except the incidence of hospitalized cases were closer to log-normal than normal distributions. The incidence of hospitalized cases was slightly closer to normal, but even under the log-normal model, observed and expected values were not substantially divergent. To maintain a consistent analytical framework, all subsequent analyses used log_10_-transformed values for wastewater viral RNA concentration, SARS-CoV-2 RNA/PMMoV RNA ratio, and the incidence of confirmed, hospitalized, and severe cases. Weekly representative values for wastewater SARS-CoV-2 RNA concentration and SARS-CoV-2 RNA/PMMoV RNA ratio were calculated as the arithmetic mean of the log_10_ values (equivalent to the geometric mean before log transformation) (Figure 1, Table S2).

**Figure 1.**
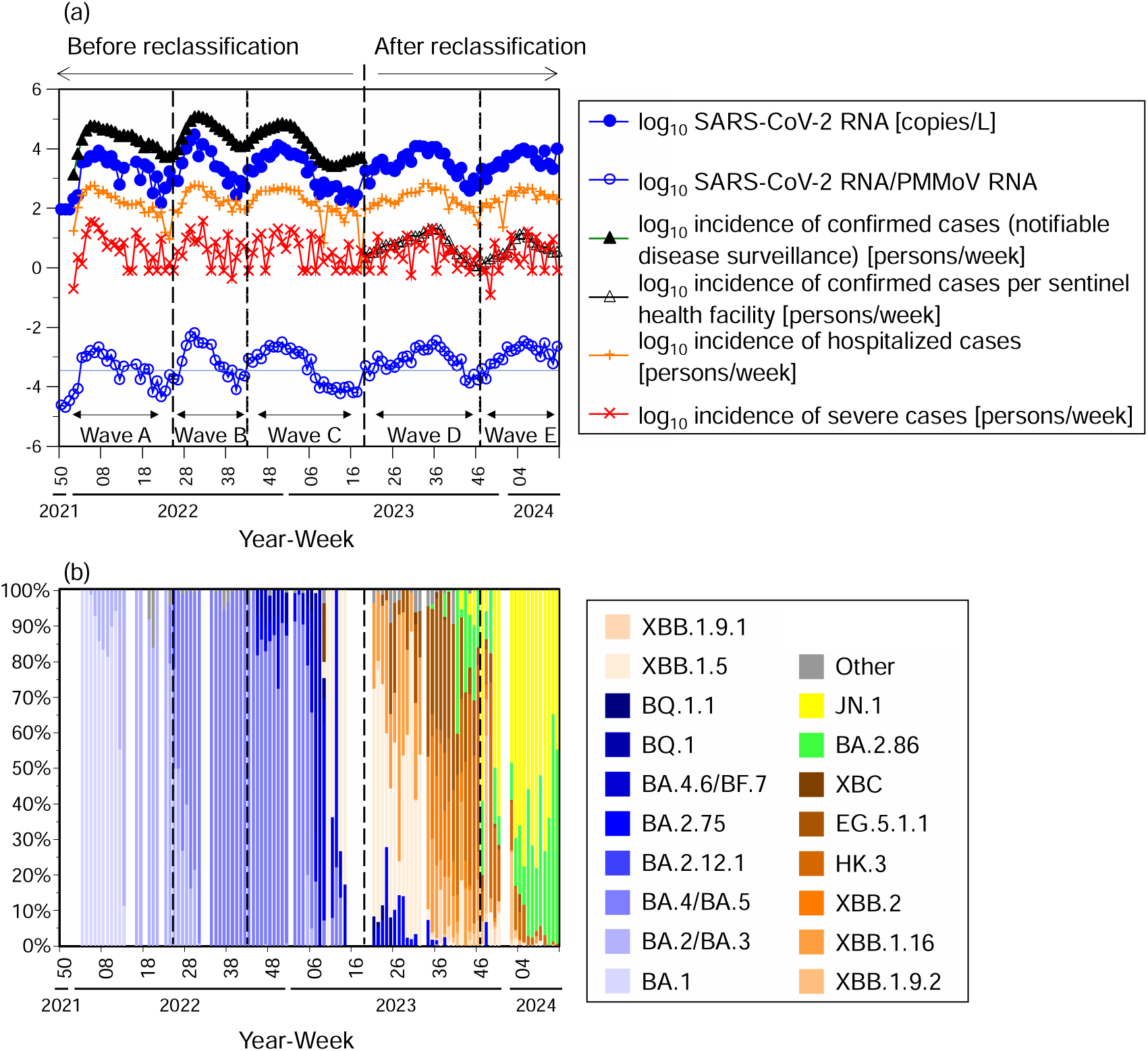
Temporal trends in SARS-CoV-2 in wastewater and COVID-19 cases. (a) SARS-CoV-2 RNA concentrations in wastewater and the incidence of confirmed, hospitalized, and severe cases; (b) SARS-CoV-2 lineage proportions in wastewater.

Hierarchical multiple regression analysis was conducted over 117 weeks from Week 1, 2022, to Week 13, 2024. Outcome variables were the incidence of confirmed, hospitalized, and severe cases. In Model 1, independent variables included same-week wastewater SARS-CoV-2 RNA concentration and wave dummy variables. Waves were defined as Wave A, weeks 1–24, 2022; Wave B, weeks 25–42, 2022; Wave C, weeks 43, 2022–18, 2023; Wave D, weeks 19–46, 2023; Wave E, weeks 47, 2023–13, 2024 (Figure 1). Wave C was the reference, as it represented the longest period immediately preceding the reclassification. Model 2 added the wastewater SARS-CoV-2 RNA concentration one week prior, Model 3 added that two weeks prior, and Model 4 added that three weeks prior. For weeks 14, 15, 33 of 2022, and week 18 of 2023, where wastewater data were missing, the previous week’s value was substituted. Regression equations are shown in equation (3):

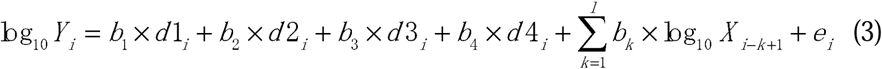

where *Y* denotes the incidence of cases, *b* denotes partial regression coefficients, *d1–4* denote wave dummy variables (Wave A, B, D, E), *l* denotes Model number, *X* denotes SARS-CoV-2 RNA concentration in wastewater, and *e* denotes the error term.

The coefficient b for each wave dummy variable represents the multiplicative change in the incidence of cases at the same wastewater RNA concentration relative to Wave C. For the incidence of confirmed cases, totals were used for pre-reclassification (Waves A–C) and per sentinel health facility for post-reclassification (Waves D–E), with units differing. Denoting the unit conversion factor as *A*, the relationship between pre- and post-reclassification cases is expressed in equation (4).

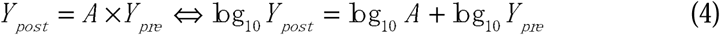

Therefore, the model for the incidence of confirmed cases is described in equation (5)

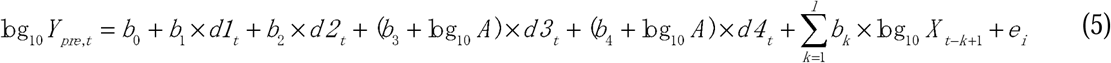

Consequently, partial regression coefficients for the wave D and E dummy variables reflect both the wave effect and the unit conversion factor.

Model selection proceeded until the change in *R²* became insignificant between successive models, with Model *k* adopted. Model validity was further evaluated using Akaike information criterion (AIC) and Bayesian information criterion (BIC). This model was designated as the main model. For hospitalized and severe cases, prevalence was also estimated from model-predicted incidence using equations (1–2), as prevalence is a practically important indicator. Sensitivity analyses considered alternative hospital stay lengths for hospitalized and severe cases, as well as models using the log_10_ SARS-CoV-2 RNA/PMMoV RNA ratio instead of wastewater SARS-CoV-2 RNA concentration.

Additional analyses used variant proportions (Figure 1, Table S3) in place of wave dummy variables, with BA.4/BA.5 as the reference. Variant values were either averaged over multiple weeks (for the incidence of confirmed cases: same week, one week prior, two weeks prior; for new hospitalized and severe cases: same week, one week prior) or used only for the week with the largest unstandardized partial regression coefficient in the main model. For weeks lacking variant data, the previous week’s value was used. For the incidence of confirmed cases, a dummy variable representing the unit conversion factor was created (pre-reclassification = 0, post-reclassification = 1).

While this model is useful for theoretical verification, practical applications require real-time estimation without waiting for a wave to end. Accordingly, data from Waves A–D were analyzed using the same variables as the main model. Predicted values for Wave E were estimated assuming it corresponded to Wave D, then corrected using the average difference between observed and predicted values from the start of Wave E to the prior week. Because case data become available one week after wastewater sampling, this correction is practically feasible. The root mean square error (RMSE) was calculated for both the parameter estimation and validation periods, as it better reflects prediction accuracy than *R²* ^29^.

## Results

Among the outcomes of confirmed, hospitalized, and severe cases, our main analyses focused on predicting the incidence of confirmed cases and the prevalence of hospitalized and severe cases. Predictions of the incidence of hospitalized and severe cases are presented in the supplemental analyses, as prevalence is more relevant than incidence for healthcare capacity planning.

For the incidence of confirmed cases, the transitions from Model 1 to Model 2 and from Model 2 to Model 3 were significant, whereas the transition from Model 3 to Model 4 was not. Therefore, Model 3 was considered appropriate (Table 1), further supported by its lowest AIC and BIC values. Similarly, Model 2 was appropriate for both the incidence of hospitalized and severe cases.

**Table 1.**
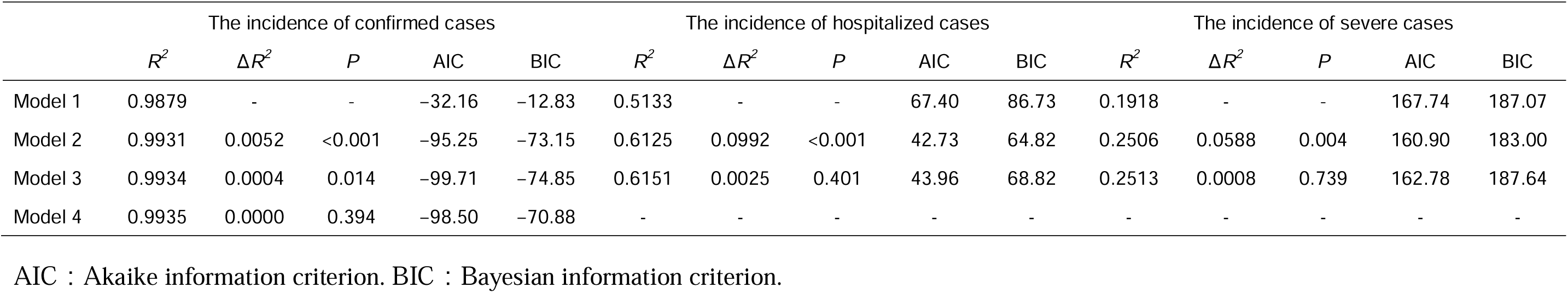
Model comparisons of hierarchical multiple regression analyses.

|  | The incidence of confirmed cases |  |  |  |  | The incidence of hospitalized cases |  |  |  |  | The incidence of severe cases |  |  |  |  |
| --- | --- | --- | --- | --- | --- | --- | --- | --- | --- | --- | --- | --- | --- | --- | --- |
| | $R^2$ | $\Delta R^2$ | $P$ | AIC | BIC | $R^2$ | $\Delta R^2$ | $P$ | AIC | BIC | $R^2$ | $\Delta R^2$ | $P$ | AIC | BIC |
| Model 1 | 0.9879 | - | - | -32.16 | -12.83 | 0.5133 | - | - | 67.40 | 86.73 | 0.1918 | - | - | 167.74 | 187.07 |
| Model 2 | 0.9931 | 0.0052 | <0.001 | -95.25 | -73.15 | 0.6125 | 0.0992 | <0.001 | 42.73 | 64.82 | 0.2506 | 0.0588 | 0.004 | 160.90 | 183.00 |
| Model 3 | 0.9934 | 0.0004 | 0.014 | -99.71 | -74.85 | 0.6151 | 0.0025 | 0.401 | 43.96 | 68.82 | 0.2513 | 0.0008 | 0.739 | 162.78 | 187.64 |
| Model 4 | 0.9935 | 0.0000 | 0.394 | -98.50 | -70.88 | - | - | - | - | - | - | - | - | - | - |

AIC : Akaike information criterion. BIC : Bayesian information criterion.

For the incidence of confirmed cases, the partial regression coefficients for same-week, one-week prior, and two-week prior wastewater SARS-CoV-2 RNA concentrations were largest in that order, and were all positive and statistically significant (Table 2). For the incidence of hospitalized cases, the coefficients for same-week and one-week prior were positive and significant, with the one-week prior coefficient slightly larger than that for the same week. For the incidence of severe cases, only the one-week prior coefficient was positive and significant, whereas the same-week coefficient was not significant.

**Table 2.**
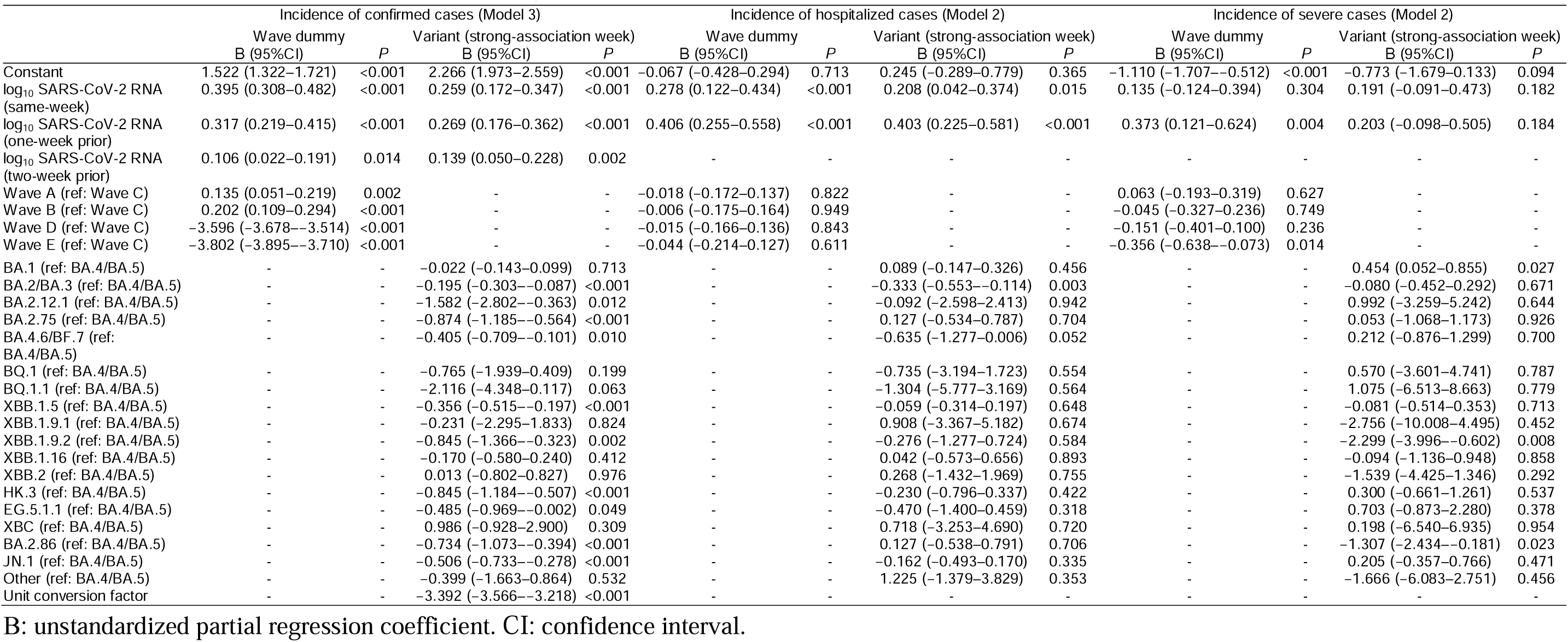
Unstandardized partial regression coefficients for the incidence of confirmed, hospitalized, and severe cases.

Regarding wave dummy variables, for the incidence of confirmed cases, Waves A and B showed significantly positive coefficients, whereas Waves D and E had significantly negative coefficients, with Wave C as the reference. No significant differences were observed for wave dummy variables in the incidence of hospitalized cases. For the incidence of severe cases, Wave E was significantly negative. Predicted values from regression analyses using wave dummy variables showed high agreement with observed values for the incidence of confirmed cases and the prevalence of hospitalized and severe cases for the overall study period (*R²* = 0.8199–0.9961; Figure 2). Associations were weaker for the incidence of hospitalized cases (*R²* = 0.6125) and the lowest for the incidence of severe cases (*R²* = 0.2506; Figure S2).

**Figure 2.**
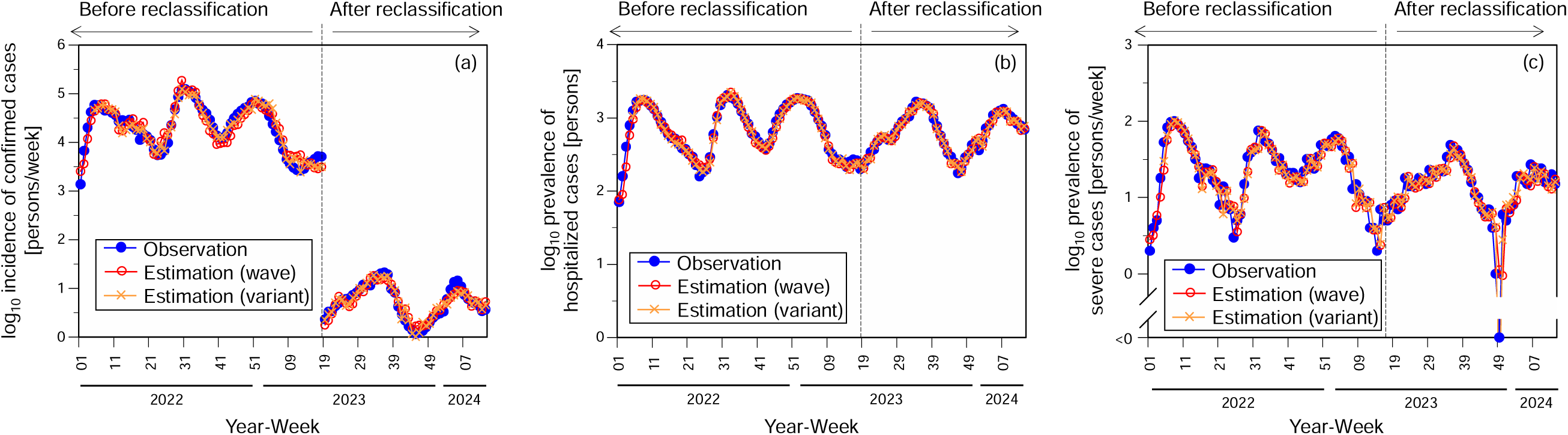
Comparison between observed and estimated values based on regression models using wave dummy and variant proportion variables. (a) Incidence of confirmed cases (wave dummy: *R^2^* = 0.9934 for overall period; 0.8851 for notifiable disease surveillance period; *R^2^* = 0.8930 for sentinel surveillance period, variant (strong-association week): *R^2^* = 0.9961 for overall period; 0.9360 for notifiable disease surveillance period; *R^2^* = 0.9030 for sentinel surveillance period); (b) prevalence of hospitalized cases (wave dummy: *R^2^* = 0.9666; variant (strong-association week): *R^2^* = 0.9795); (c) prevalence of severe cases (wave dummy: *R^2^* = 0.8199; variant (strong-association week): *R^2^* = 0.8451 [the calculation excludes data from one week when the number of severe cases was zero]).

Sensitivity analyses that varied hospital stay length indicated that the magnitudes of partial regression coefficients for wastewater concentrations remained largely consistent with the main model results. However, unlike the main model, wave dummy variables showed significant associations with the incidence of hospitalized cases (Tables S4, S5). Nevertheless, the incidence of severe cases consistently exhibited stronger associations with wave variables than hospitalized cases. Adjusting for PMMoV yielded similar results, with no substantial improvement in model fit (Tables S6, Figure S3).

Regression analyses using variant proportions showed that models employing the average variant proportion across multiple weeks provided a slightly better fit for the incidence of confirmed cases than models using single-week data (Table S7). Conversely, models using single-week variant proportions better fit for the incidence of hospitalized and severe cases. Using single-week variant proportions, the incidence of confirmed cases was significantly associated with various variants, whereas the incidence of hospitalized cases was significantly associated only with BA.2/BA.3, with BA.4/BA.5 as the reference. For the incidence of severe cases, BA.1 showed a positive association, whereas XBB.1.9.2 and BA.2.86 were significantly negatively associated. The dummy variable for the unit conversion factor had a value of −3.392.

Comparisons between predicted and observed values, using variant proportions, showed good agreement for the incidence of confirmed and hospitalized cases and for the prevalence of hospitalized and severe cases, except for the incidence of severe cases, which was similar to the observed values when wave dummy variables were included (Figures 2 and S2).

Regression analyses of Waves A–D produced similar results, except that the wastewater RNA concentration one week prior showed a borderline significant association with the incidence of severe cases (*P* = 0.055). While agreement was somewhat lower for the incidence of hospitalized cases (RMSE = 0.2641) and the incidence of severe cases (RMSE = 0.5486) during the validation period, RMSE values for the incidence of confirmed cases and the prevalence of hospitalized and severe cases indicated good agreement, ranging from 0.0665 to 0.2065 (Figures 3, S4, Table S8).

**Figure 3.**
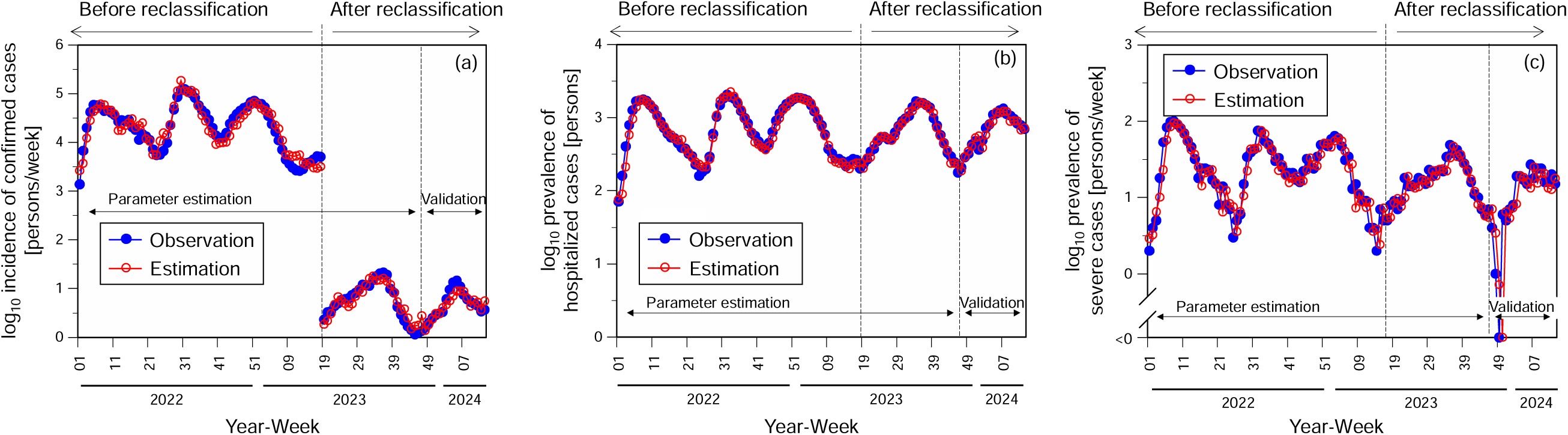
Comparison between observed and estimated values based on regression models fitted to Waves A–D with calibration using data from the validation period (Wave E). (a) Incidence of confirmed cases (root mean squared error [RMSE] = 0.1521 for overall parameter estimation period; 0.1626 for notifiable disease surveillance at parameter estimation period; RMSE = 0.1219 for sentinel surveillance at parameter estimation period; RMSE = 0.1392 at validation period); (b) prevalence of hospitalized cases (RMSE = 0.0618 at parameter estimation period; RMSE = 0.0665 at validation period); (c) prevalence of severe cases (RMSE = 0.1434 at parameter estimation period; RMSE = 0.2065 at validation period [the calculation excludes data from two weeks when the number of severe cases was zero]).

## Discussion

This study shows that SARS-CoV-2 RNA concentrations in wastewater can predict the incidence of COVID-19 confirmed cases and the prevalence of hospitalized and severe cases, even without PMMoV normalization. The high predictive accuracy of the models (*R²* = 0.8199–0.9961) underscores the potential of wastewater surveillance to provide early signals of disease burden. By applying wave E corrections to regression models derived from Waves A–D, the RMSE during the validation period remained sufficiently low (0.0665–0.2065), demonstrating good predictive performance. The low to moderate agreement for the incidence of hospitalized and severe cases likely reflects high variability in converting prevalence to incidence. Nevertheless, this study was able to accurately estimate the prevalence of hospitalized and severe cases of practical clinical significance through wastewater surveillance. Because wastewater viral RNA concentrations can be measured approximately 1–2 days after sampling, the results can theoretically be obtained about one week earlier than the published confirmed, hospitalized, and severe case data. These findings therefore underscore the practical value of wastewater surveillance. Overall, monitoring SARS-CoV-2 RNA concentrations in wastewater enables prediction of the incidence of confirmed cases and the prevalence of hospitalized and severe cases approximately one week in advance, providing useful information for healthcare capacity planning.

The incidence of confirmed cases was associated with wastewater viral RNA concentrations in the same week, one week prior, and two weeks prior, with the strength of association decreasing with increasing lag. This suggests that a weighted average incorporating three weeks of wastewater viral RNA concentration data will likely reduce variability by integrating multi-week data, as previously reported ^10^. The particularly strong correlation with the same week is consistent with preceding studies indicating that the diagnostic lag from wastewater viral RNA detection to clinically confirmed cases is at most a few days and that same-week correlations are strongest ^6,11^. In contrast, the incidence of hospitalized and severe cases showed stronger correlations with wastewater viral RNA concentrations one week prior than with those from the same week, consistent with earlier findings ^7,8^. These correlations are consistent with the typical clinical course in which hospitalization and severe illness occur after symptom onset ^30^.

Importantly, the incidence of confirmed and severe cases was significantly associated with the wave dummy variables, whereas the incidence of hospitalized cases was not. For the incidence of confirmed cases, Waves A–C were based on total case counts, whereas Waves D–E used sentinel surveillance data; therefore, the lower partial regression coefficients for Waves D and E are expected a priori. Moreover, the partial regression coefficients for Waves A and B were significantly positive, and the coefficient for Wave E was smaller than that for Wave D, with Wave C as the reference. The wave dummy coefficients, which represent time-specific effects after adjusting for wastewater SARS-CoV-2 RNA concentrations, decreased over successive waves, indicating a decrease in the number of cases per unit of wastewater viral RNA concentration.

A comparison of the model including wave dummy variables and that including variant proportion variables showed that the partial regression coefficients were 0.2 and 0.4 lower for Waves D and E, respectively, than those based on the unit conversion factor (bottom row in Table 2). This corresponds to an approximate 40% (= 1 − 10^−0.2^) and 60% (= 1 − 10^−0.4^) reduction in the incidence of confirmed cases relative to wastewater RNA concentrations compared with Wave C, consistent with a previously reported 50% decrease after reclassification ^10^. The partial regression coefficient for the incidence of severe cases was also significantly smaller in Wave E. Taken together, these findings indicate that the ratio of the incidence of confirmed and severe cases to wastewater viral RNA concentration decreased over time, whereas the ratio for hospitalized cases remained relatively stable across five waves analyzed. Sensitivity analyses varying length of hospital stay showed significant associations between the incidence of hospitalized cases and wave dummy variables in some models; however, the association was consistently stronger for severe cases than for hospitalized cases.

Among the four potential contributors to divergence between wastewater surveillance and reported case counts over time—specifically, (1) increased viral load excreted by an infected individual, (2) reduced patients’ healthcare-seeking, (3) decreased testing at medical institutions, and (4) milder symptoms due to virulence attenuation or accumulated immunity, factor (1) is unlikely to be the primary explanation. If increased viral shedding were the dominant factor, the incidence of hospitalized cases would also show similar wave associations. This interpretation is consistent with previous studies reporting that post-Omicron fecal viral RNA concentrations did not vary substantially across most variants, except BA.2.86 and JN.1 ^18^.

In contrast, the declining trend in the incidence of confirmed and severe cases relative to wastewater viral RNA concentrations suggests contributions from factors (2), (3), and (4). Regarding the incidence of confirmed cases, reduced healthcare-seeking and reduced testing at medical facilities have been previously reported ^13,14^. This trend was particularly evident after reclassification, when public subsidies for COVID-19-related test and hospitalization costs were terminated. However, even prior to reclassification, the reduction in insurance benefit payments for home or lodging-based recuperation since September 2022 had potentially contributed.

The milder clinical presentation hypothesis is supported by the pronounced decline in the incidence of severe cases during Wave E. Variant-proportion analyses showed associations between emerging lineages from Waves A to E and the incidence of confirmed cases. In contrast, for the incidence of severe cases, BA.1 (dominant in Wave A) showed a positive association compared with BA.4/BA.5 as the reference, whereas XBB.1.9.2 and BA.2.86 (emerging in Wave E) showed significant negative associations. Unlike the incidence of confirmed cases, that of severe cases is likely to be less sensitive to reduced healthcare-seeking and testing. Therefore, the decline in the incidence of severe cases during Wave E may largely reflect reduced virulence of XBB.1.9.2 and BA.2.86, although the contribution of accumulated immunity cannot be excluded. Taken together, while susceptibility to hospitalization (hospitalizations per infected individual) remained relatively stable, reductions in healthcare-seeking for mild symptoms, decreased medical testing, and lower probability of progression to severe illness appear to have occurred over time. These findings suggest that wastewater surveillance can provide a robust indicator of disease burden even when clinical surveillance is affected by changes in healthcare-seeking behavior or testing practices.

This study has several limitations. First, generalizability may be limited due to the restricted geographic scope of the study areas. Second, the analysis period may also limit generalizability, covering from January 2022 to March 2024 without accounting for subsequent waves or variant-related changes (e.g., NB.1.8.1 lineage). Third, Japan modified its surveillance system for hospitalized and severe cases after April 2024, requiring the development of new predictive models under the updated framework.

Despite these limitations, this study demonstrates that wastewater surveillance can predict the incidence of confirmed cases and the prevalence of hospitalized and severe cases in advance, thus supporting healthcare capacity planning. It also provides insights into the viral, clinical, and surveillance-related mechanisms underlying divergence between wastewater data and reported case counts, including reduced patients’ healthcare-seeking, decreased medical testing, and milder clinical manifestations associated with variant evolution and accumulated immunity.

## Data Availability

SARS-CoV-2 RNA and PMMoV RNA concentrations in wastewater are available at https://www.pref.kanagawa.jp/docs/h2d/covid19/simulation.html. Other data are available from the corresponding author, Byung-Kwang Yoo, upon reasonable request.

## Contributors

MM, RW, UC, MK, and BKY contributed to conceptualization.

MM contributed to methodology, formal analysis, and visualization.

RI contributed to investigation.

BKY contributed to supervision.

MM and BKY contributed to funding acquisition.

MM and RI contributed to writing – original draft.

All authors contributed to data interpretation, critically revised the manuscript, and approved the final version for submission.

## Acknowledgments

This study was supported by The Kanagawa University of Human Services and The Nippon Foundation–The University of Osaka Project for Infectious Disease Prevention. The funders had no involvement in the study design, data collection, data analysis, data interpretation, or manuscript preparation.

## AI Statements

The authors used DeepL and ChatGPT to refine the English language of the manuscript. The manuscript was originally written by the authors, who critically reviewed and edited all the AI-assisted revisions. The authors take full responsibility for the content of this publication.

## References

1. World Health Organization. WHO COVID-19 dashboard. 2026. https://data.who.int/dashboards/covid19/ (accessed February 19, 2026).

2. U.S. Centers for Disease Control and Prevention. Provisional COVID-19 mortality surveillance. 2026. https://www.cdc.gov/nchs/nvss/vsrr/covid19/ (accessed February 19, 2026).

3. Ministory of Health Labour and Welfare. Vital statistics. 2026. https://www.e-stat.go.jp/en/stat-search/files?page=1&toukei=00450011&tstat=000001028897 (accessed February 19, 2026).

4. Kimura H, Hosozawa M, Taniguchi Y, et al. COVID-19-specific prefectural hospital bed utilization rate and in-hospital mortality among COVID-19 patients throughout the first 3 years of the pandemic in Japan. J Epidemiol 2025; 35(9): 402–9.

5. Shah S, Gwee SXW, Ng JQX, Lau N, Koh J, Pang J. Wastewater surveillance to infer COVID-19 transmission: A systematic review. Sci Total Environ 2022; 804: 150060.

6. Ando H, Murakami M, Ahmed W, Iwamoto R, Okabe S, Kitajima M. Wastewater-based prediction of COVID-19 cases using a highly sensitive SARS-CoV-2 RNA detection method combined with mathematical modeling. Environ Int 2023; 173: 107743.

7. Galani A, Aalizadeh R, Kostakis M, et al. SARS-CoV-2 wastewater surveillance data can predict hospitalizations and ICU admissions. Sci Total Environ 2022; 804: 150151.

8. Hegazy N, Cowan A, D’Aoust PM, et al. Understanding the dynamic relation between wastewater SARS-CoV-2 signal and clinical metrics throughout the pandemic. Sci Total Environ 2022; 853: 158458.

9. Boehm AB, Wolfe MK, White B, Hughes B, Duong D. Divergence of wastewater SARS-CoV-2 and reported laboratory-confirmed COVID-19 incident case data coincident with wide-spread availability of at-home COVID-19 antigen tests. PeerJ 2023; 11: e15631.

10. Murakami M, Ando H, Yamaguchi R, Kitajima M. Evaluating survey techniques in wastewater-based epidemiology for accurate COVID-19 incidence estimation. Sci Total Environ 2024; 954: 176702.

11. Murakami M, Ishiguro N, Ando H, et al. Insights from wastewater surveillance into testing-related underreporting and hospital-acquired SARS-CoV-2 infections. Environ Int 2026; 207: 110028.

12. Daou M, Kannout H, Khalili M, et al. Analysis of SARS-CoV-2 viral loads in stool samples and nasopharyngeal swabs from COVID-19 patients in the United Arab Emirates. PLoS ONE 2022; 17(9): e0274961.

13. Maree G, Els F, Naidoo Y, et al. Wastewater surveillance overcomes socio-economic limitations of laboratory-based surveillance when monitoring disease transmission: The South African experience during the COVID-19 pandemic. PLoS ONE 2025; 20(2): e0311332.

14. Makino M, Takesue Y, Murakami Y, et al. Influence of easing COVID-19 strategies following downgrading of the national infectious disease category on COVID-19 occurrence among hospitalized patients in Japan. J Infect Chemother 2025; 31(2): 102464.

15. Arabi M, Al-Najjar Y, Mhaimeed N, et al. Severity of the Omicron SARS-CoV-2 variant compared with the previous lineages: A systematic review. J Cell Mol Med 2023; 27(11): 1443–64.

16. Kanagawa Prefectural Government. COVID-19 data archive (translated by authors). 2025. https://www.pref.kanagawa.jp/docs/ga4/covid19/archive/data.html (accessed February 19, 2026). (in Japanese)

17. Japan Institute for Health Security. IDWR surveillance data table. 2026. https://id-info.jihs.go.jp/en/surveillance/idwr/rapid/ (accessed February 19, 2026).

18. Wannigama DL, Amarasiri M, Phattharapornjaroen P, et al. Increased faecal shedding in SARS-CoV-2 variants BA.2.86 and JN.1. Lancet Infect Dis 2024; 24(6): e348–e50.

19. Li X, Zhang S, Sherchan S, et al. Correlation between SARS-CoV-2 RNA concentration in wastewater and COVID-19 cases in community: A systematic review and meta-analysis. J Hazard Mater 2023; 441: 129848.

20. Gordis L. Epidemiology. Fifth Edition. Philadephia, United States: Elsevier 2014.

21. Andersen KM, Brouillette MA, Togo K, et al. Inpatient burden of COVID-19 in Japan: A retrospective cohort study. J Infect Chemother 2025; 31(7): 102721.

22. Uno S, Goto R, Honda K, et al. Healthcare costs for hospitalized COVID-19 patients in a Japanese university hospital: a cross-sectional study. Cost Eff Resour Alloc 2023; 21(1): 43.

23. Matsunaga N, Hayakawa K, Terada M, et al. Clinical epidemiology of hospitalized patients with coronavirus disease 2019 (COVID-19) in Japan: Report of the COVID-19 registry Japan. Clin Infect Dis 2020; 73(11): e3677–e89.

24. Ando H, Iwamoto R, Kobayashi H, Okabe S, Kitajima M. The Efficient and Practical virus Identification System with ENhanced Sensitivity for Solids (EPISENS-S): A rapid and cost-effective SARS-CoV-2 RNA detection method for routine wastewater surveillance. Sci Total Environ 2022; 843: 157101.

25. Kuroiwa M, Gahara Y, Kato H, et al. Targeted amplicon sequencing of wastewater samples for detecting SARS-CoV-2 variants with high sensitivity and resolution. Sci Total Environ 2023; 893: 164766.

26. Lee L. Package ’NADA’. 2022. https://cran.r-project.org/web/packages/NADA/NADA.pdf (accessed November 14, 2025).

27. R Development Core Team. R 4.5.0. R: A language and environment for statistical computing. Vienna, Austria 2025.

28. Mishra P, Pandey CM, Singh U, Gupta A, Sahu C, Keshri A. Descriptive statistics and normality tests for statistical data. Ann Card Anaesth 2019; 22(1): 67–72.

29. Kuhn M, Johnson K. Measuring Performance in Regression Models. In: Kuhn M, Johnson K, eds. Applied Predictive Modeling. New York, NY: Springer New York; 2013: 95-100.

30. Cevik M, Bamford CGG, Ho A. COVID-19 pandemic—a focused review for clinicians. Clin Microbiol Infect 2020; 26(7): 842–7.

