## Supplementary Materials for "Prediction of confirmed, hospitalized, and severe COVID-19 cases and mechanistic insights from viral concentrations and variant dynamics in wastewater"

Table S1. Value corresponding to half of the ND (non-detect) proportion. CI: confidence interval.

| Items | Distribution | Estimated mean (95% CI) | Value corresponding to half of the ND proportion |
|---|---|---|---|
| SARS-CoV-2 RNA concentrations in wastewater [copies/L] | Normal distribution | 5640 (4683–6596) | −10655 |
| SARS-CoV-2 RNA concentrations in wastewater [copies/L] | Log-normal distribution | 2007 (1708–2358) | 129 |
| Incidence of hospitalized cases [persons/week] | Normal distribution | 234.2 (204.4–264.0) | −193.4 |
| Incidence of hospitalized cases [persons/week] | Log-normal distribution | 166.9 (138.9–200.5) | 11.93 |
| Incidence of severe cases [persons/week] | Normal distribution | 6.08 (4.76–7.41) | −3.07 |
| Incidence of severe cases [persons/week] | Log-normal distribution | 3.40 (2.76–4.19) | 0.80 |

Table S2. SARS-CoV-2 RNA concentrations, SARS-CoV-2 RNA/PMMoV RNA ratios in wastewater, the incidence of confirmed cases, and the prevalence of hospitalized and severe cases for each week.

| Year-Week | Number of wastewater samples | SARS-CoV-2 RNA [copies/L] | SARS-CoV-2 RNA/PMMoV RNA [$\times10^{-6}$] | Incidence of confirmed cases (notifiable disease surveillance) [persons/week] | Incidence of confirmed cases per sentinel health facility [persons/week] | Prevalence of hospitalized cases [persons] | Prevalence of severe cases [persons] |
|---|---|---|---|---|---|---|---|
| 202147 | - | - | - | - | - | - | 5 |
| 202148 | - | - | - | - | - | 35 | 2 |
| 202149 | - | - | - | - | - | 18 | 0 |
| 202150 | 6 | 93 | 25 | - | - | 35 | 0 |
| 202151 | 6 | 93 | 21 | - | - | 62 | 1 |
| 202152 | 2 | 93 | 34 | - | - | 65 | 2 |
| 202201 | 4 | 208 | 58 | 1379 | - | 71 | 2 |
| 202202 | 4 | 269 | 88 | 6738 | - | 161 | 4 |
| 202203 | 6 | 3470 | 951 | 20082 | - | 406 | 5 |
| 202204 | 6 | 3735 | 1086 | 41838 | - | 795 | 18 |
| 202205 | 6 | 5483 | 1657 | 58794 | - | 1262 | 53 |
| 202206 | 4 | 5373 | 1393 | 57875 | - | 1668 | 83 |
| 202207 | 6 | 8526 | 2225 | 53327 | - | 1726 | 98 |
| 202208 | 4 | 5724 | 1261 | 44728 | - | 1778 | 100 |
| 202209 | 6 | 3579 | 730 | 45609 | - | 1688 | 91 |
| 202210 | 6 | 5540 | 1208 | 40815 | - | 1486 | 81 |
| 202211 | 6 | 2888 | 549 | 36362 | - | 1320 | 69 |
| 202212 | 4 | 618 | 178 | 26777 | - | 1071 | 55 |
| 202213 | 4 | 2292 | 493 | 28779 | - | 886 | 46 |
| 202214 | 0 | - | - | 27217 | - | 716 | 32 |
| 202215 | 0 | - | - | 28836 | - | 603 | 18 |
| 202216 | 4 | 3607 | 581 | 20760 | - | 532 | 24 |
| 202217 | 2 | 882 | 179 | 17984 | - | 522 | 24 |
| 202218 | 2 | 3007 | 402 | 11282 | - | 446 | 23 |
| 202219 | 4 | 2346 | 391 | 14698 | - | 388 | 17 |
| 202220 | 4 | 321 | 67 | 13263 | - | 379 | 15 |
| 202221 | 4 | 1157 | 160 | 11208 | - | 320 | 8 |
| 202222 | 4 | 151 | 47 | 7190 | - | 298 | 14 |
| 202223 | 4 | 480 | 72 | 5504 | - | 227 | 8 |
| 202224 | 4 | 1720 | 258 | 5598 | - | 160 | 7 |
| 202225 | 4 | 849 | 191 | 6705 | - | 183 | 3 |
| 202226 | 4 | 833 | 173 | 9742 | - | 199 | 5 |
| 202227 | 4 | 3302 | 725 | 22658 | - | 293 | 6 |

| | | | | | | | |
|---|---|---|---|---|---|---|---|
| 202228 | 4 | 10245 | 2474 | 47295 | - | 599 | 15 |
| 202229 | 4 | 19101 | 4955 | 86193 | - | 1032 | 34 |
| 202230 | 4 | 29530 | 6620 | 117822 | - | 1495 | 39 |
| 202231 | 4 | 5597 | 3080 | 124535 | - | 1849 | 43 |
| 202232 | 2 | 14044 | 2977 | 112680 | - | 2064 | 75 |
| 202233 | 0 | - | - | 102046 | - | 1904 | 71 |
| 202234 | 4 | 8315 | 2669 | 82177 | - | 1820 | 55 |
| 202235 | 4 | 2521 | 789 | 58055 | - | 1568 | 51 |
| 202236 | 4 | 2363 | 500 | 45930 | - | 1230 | 42 |
| 202237 | 4 | 1794 | 462 | 39502 | - | 991 | 30 |
| 202238 | 2 | 683 | 213 | 29243 | - | 830 | 33 |
| 202239 | 4 | 1122 | 369 | 20048 | - | 619 | 25 |
| 202240 | 4 | 280 | 80 | 13910 | - | 564 | 20 |
| 202241 | 4 | 834 | 264 | 12374 | - | 471 | 21 |
| 202242 | 4 | 546 | 229 | 12748 | - | 404 | 21 |
| 202243 | 4 | 1927 | 891 | 15599 | - | 380 | 17 |
| 202244 | 4 | 1711 | 676 | 24317 | - | 451 | 16 |
| 202245 | 4 | 4571 | 1209 | 31357 | - | 640 | 22 |
| 202246 | 4 | 2439 | 880 | 38283 | - | 905 | 32 |
| 202247 | 4 | 6510 | 1743 | 44529 | - | 1140 | 25 |
| 202248 | 4 | 5509 | 2470 | 50369 | - | 1333 | 24 |
| 202249 | 4 | 7755 | 2145 | 54166 | - | 1543 | 34 |
| 202250 | 4 | 13242 | 2198 | 67035 | - | 1635 | 49 |
| 202251 | 4 | 10857 | 3202 | 71467 | - | 1782 | 50 |
| 202252 | 2 | 8892 | 1801 | 62081 | - | 1873 | 47 |
| 202301 | 2 | 6393 | 2030 | 61779 | - | 1855 | 58 |
| 202302 | 4 | 6835 | 1308 | 52256 | - | 1803 | 64 |
| 202303 | 4 | 5700 | 1574 | 35098 | - | 1733 | 60 |
| 202304 | 4 | 5048 | 1288 | 23707 | - | 1481 | 47 |
| 202305 | 4 | 1611 | 377 | 16508 | - | 1211 | 31 |
| 202306 | 4 | 2588 | 865 | 11000 | - | 952 | 34 |
| 202307 | 4 | 613 | 198 | 7124 | - | 784 | 13 |
| 202308 | 4 | 299 | 91 | 4468 | - | 586 | 15 |
| 202309 | 4 | 716 | 150 | 3638 | - | 372 | 11 |
| 202310 | 4 | 378 | 92 | 3014 | - | 324 | 9 |
| 202311 | 4 | 516 | 82 | 2689 | - | 273 | 9 |
| 202312 | 2 | 551 | 97 | 2626 | - | 273 | 4 |
| 202313 | 4 | 193 | 59 | 2863 | - | 253 | 4 |
| 202314 | 4 | 327 | 96 | 3774 | - | 217 | 2 |
| 202315 | 4 | 361 | 102 | 3944 | - | 253 | 7 |

| | | | | | | | |
|---|---|---|---|---|---|---|---|
| 202316 | 4 | 162 | 64 | 4514 | - | 272 | 5 |
| 202317 | 2 | 275 | 67 | 5182 | - | 265 | 5 |
| 202318 | 0 | - | - | 5068 | - | 202 | 8 |
| 202319 | 4 | 1850 | 528 | - | 2.33 | 232 | 9 |
| 202320 | 4 | 683 | 236 | - | 3.30 | 268 | 7 |
| 202321 | 4 | 2007 | 426 | - | 3.43 | 342 | 9 |
| 202322 | 4 | 3474 | 1075 | - | 4.38 | 373 | 18 |
| 202323 | 4 | 4230 | 729 | - | 5.03 | 480 | 14 |
| 202324 | 4 | 2105 | 394 | - | 5.91 | 540 | 14 |
| 202325 | 4 | 2288 | 546 | - | 6.07 | 545 | 15 |
| 202326 | 4 | 1794 | 450 | - | 6.30 | 508 | 18 |
| 202327 | 4 | 2858 | 638 | - | 7.38 | 505 | 17 |
| 202328 | 4 | 5269 | 996 | - | 8.14 | 624 | 15 |
| 202329 | 4 | 6468 | 1095 | - | 9.50 | 762 | 23 |
| 202330 | 4 | 3631 | 653 | - | 12.04 | 896 | 19 |
| 202331 | 4 | 12174 | 2103 | - | 11.70 | 976 | 22 |
| 202332 | 2 | 12132 | 2435 | - | 11.55 | 1053 | 23 |
| 202333 | 2 | 11822 | 1686 | - | 15.06 | 1398 | 22 |
| 202334 | 4 | 7178 | 1818 | - | 18.10 | 1672 | 34 |
| 202335 | 4 | 11117 | 2653 | - | 20.33 | 1595 | 49 |
| 202336 | 4 | 11195 | 3623 | - | 21.43 | 1584 | 38 |
| 202337 | 4 | 7132 | 2176 | - | 19.26 | 1478 | 42 |
| 202338 | 2 | 6720 | 1576 | - | 9.85 | 1378 | 34 |
| 202339 | 4 | 2972 | 1107 | - | 8.30 | 978 | 28 |
| 202340 | 4 | 1933 | 754 | - | 4.26 | 780 | 23 |
| 202341 | 4 | 1245 | 509 | - | 2.95 | 580 | 16 |
| 202342 | 4 | 2252 | 854 | - | 2.19 | 447 | 11 |
| 202343 | 4 | 545 | 164 | - | 1.67 | 333 | 10 |
| 202344 | 2 | 390 | 134 | - | 1.41 | 296 | 7 |
| 202345 | 4 | 990 | 263 | - | 1.16 | 248 | 6 |
| 202346 | 4 | 590 | 170 | - | 1.24 | 178 | 7 |
| 202347 | 4 | 2074 | 415 | - | 1.36 | 208 | 4 |
| 202348 | 4 | 976 | 182 | - | 1.60 | 288 | 1 |
| 202349 | 4 | 2080 | 592 | - | 2.01 | 319 | 0 |
| 202350 | 4 | 2449 | 609 | - | 2.50 | 430 | 6 |
| 202351 | 4 | 3442 | 943 | - | 2.88 | 477 | 5 |
| 202352 | 2 | 2586 | 809 | - | 3.11 | 364 | 7 |
| 202401 | 2 | 5871 | 2204 | - | 3.34 | 487 | 8 |
| 202402 | 4 | 5228 | 1598 | - | 6.05 | 723 | 19 |
| 202403 | 4 | 6637 | 1804 | - | 9.57 | 808 | 19 |

| | | | | | | | |
|---|---|---|---|---|---|---|---|
| 202404 | 4 | 8730 | 2263 | - | 13.56 | 1097 | 17 |
| 202405 | 4 | 10143 | 3452 | - | 14.61 | 1130 | 15 |
| 202406 | 4 | 7800 | 2819 | - | 11.01 | 1265 | 27 |
| 202407 | 4 | 8142 | 2345 | - | 7.41 | 1329 | 18 |
| 202408 | 2 | 3970 | 1687 | - | 6.10 | 1114 | 24 |
| 202409 | 4 | 2699 | 1031 | - | 5.33 | 1066 | 24 |
| 202410 | 4 | 8460 | 2983 | - | 4.93 | 963 | 16 |
| 202411 | 4 | 3109 | 1258 | - | 4.45 | 828 | 16 |
| 202412 | 4 | 2157 | 601 | - | 3.43 | 787 | 20 |
| 202413 | 2 | 10039 | 2337 | - | 3.69 | 696 | 15 |

Table S3. Proportion of SARS-CoV-2 lineages in wastewater for each week.

| Year-Week | N | BA.1 | BA.2/BA.3 | BA.4/BA.5 | BA.2.12.1 | BA.2.75 | BA.4.6/BF.7 | BQ.1 | BQ.1.1 | XBB.1.5 | XBB.1.9.1 | XBB.1.9.2 | XBB.1.16 | XBB.2 | HK.3 | EG5.1.1 | XBC | BA.2.86 | JN.1 | Other |
|---|---|---|---|---|---|---|---|---|---|---|---|---|---|---|---|---|---|---|---|---|
| 202203 | 1 | 100% | 0% | 0% | 0% | 0% | 0% | 0% | 0% | 0% | 0% | 0% | 0% | 0% | 0% | 0% | 0% | 0% | 0% | 0% |
| 202204 | 2 | 100% | 0% | 0% | 0% | 0% | 0% | 0% | 0% | 0% | 0% | 0% | 0% | 0% | 0% | 0% | 0% | 0% | 0% | 0% |
| 202205 | 2 | 99% | 1% | 0% | 0% | 0% | 0% | 0% | 0% | 0% | 0% | 0% | 0% | 0% | 0% | 0% | 0% | 0% | 0% | 0% |
| 202206 | 2 | 93% | 7% | 0% | 0% | 0% | 0% | 0% | 0% | 0% | 0% | 0% | 0% | 0% | 0% | 0% | 0% | 0% | 0% | 0% |
| 202207 | 2 | 86% | 14% | 0% | 0% | 0% | 0% | 0% | 0% | 0% | 0% | 0% | 0% | 0% | 0% | 0% | 0% | 0% | 0% | 0% |
| 202208 | 2 | 84% | 17% | 0% | 0% | 0% | 0% | 0% | 0% | 0% | 0% | 0% | 0% | 0% | 0% | 0% | 0% | 0% | 0% | 0% |
| 202209 | 2 | 82% | 19% | 0% | 0% | 0% | 0% | 0% | 0% | 0% | 0% | 0% | 0% | 0% | 0% | 0% | 0% | 0% | 0% | 0% |
| 202210 | 2 | 90% | 10% | 0% | 0% | 0% | 0% | 0% | 0% | 0% | 0% | 0% | 0% | 0% | 0% | 0% | 0% | 0% | 0% | 0% |
| 202211 | 2 | 95% | 6% | 0% | 0% | 0% | 0% | 0% | 0% | 0% | 0% | 0% | 0% | 0% | 0% | 0% | 0% | 0% | 0% | 0% |
| 202212 | 2 | 56% | 45% | 0% | 0% | 0% | 0% | 0% | 0% | 0% | 0% | 0% | 0% | 0% | 0% | 0% | 0% | 0% | 0% | 0% |
| 202213 | 2 | 12% | 89% | 0% | 0% | 0% | 0% | 0% | 0% | 0% | 0% | 0% | 0% | 0% | 0% | 0% | 0% | 0% | 0% | 0% |
| 202214 | 0 | - | - | - | - | - | - | - | - | - | - | - | - | - | - | - | - | - | - | - |
| 202215 | 0 | - | - | - | - | - | - | - | - | - | - | - | - | - | - | - | - | - | - | - |
| 202216 | 2 | 1% | 99% | 0% | 0% | 0% | 0% | 0% | 0% | 0% | 0% | 0% | 0% | 0% | 0% | 0% | 0% | 0% | 0% | 0% |
| 202217 | 2 | 0% | 100% | 0% | 0% | 0% | 0% | 0% | 0% | 0% | 0% | 0% | 0% | 0% | 0% | 0% | 0% | 0% | 0% | 0% |
| 202218 | 0 | - | - | - | - | - | - | - | - | - | - | - | - | - | - | - | - | - | - | - |
| 202219 | 2 | 0% | 92% | 0% | 0% | 0% | 0% | 0% | 0% | 0% | 0% | 0% | 0% | 0% | 0% | 0% | 0% | 0% | 0% | 8% |
| 202220 | 1 | 0% | 84% | 0% | 0% | 0% | 0% | 0% | 0% | 0% | 0% | 0% | 0% | 0% | 0% | 0% | 0% | 0% | 0% | 16% |
| 202221 | 1 | 0% | 100% | 0% | 0% | 0% | 0% | 0% | 0% | 0% | 0% | 0% | 0% | 0% | 0% | 0% | 0% | 0% | 0% | 0% |
| 202222 | 0 | - | - | - | - | - | - | - | - | - | - | - | - | - | - | - | - | - | - | - |
| 202223 | 1 | 0% | 100% | 0% | 0% | 0% | 0% | 0% | 0% | 0% | 0% | 0% | 0% | 0% | 0% | 0% | 0% | 0% | 0% | 0% |
| 202224 | 2 | 0% | 80% | 16% | 0% | 0% | 0% | 0% | 0% | 0% | 0% | 0% | 0% | 0% | 0% | 0% | 0% | 0% | 0% | 4% |
| 202225 | 2 | 0% | 50% | 28% | 22% | 0% | 0% | 0% | 0% | 0% | 0% | 0% | 0% | 0% | 0% | 0% | 0% | 0% | 0% | 0% |
| 202226 | 2 | 0% | 50% | 50% | 0% | 0% | 0% | 0% | 0% | 0% | 0% | 0% | 0% | 0% | 0% | 0% | 0% | 0% | 0% | 0% |

| | | | | | | | | | | | | | | | | | | | | |
|---|---|---|---|---|---|---|---|---|---|---|---|---|---|---|---|---|---|---|---|---|
| 202227 | 2 | 0% | 27% | 72% | 0% | 0% | 0% | 0% | 0% | 0% | 0% | 0% | 0% | 0% | 0% | 0% | 0% | 0% | 0% | 1% |
| 202228 | 2 | 0% | 40% | 60% | 0% | 0% | 0% | 0% | 0% | 0% | 0% | 0% | 0% | 0% | 0% | 0% | 0% | 0% | 0% | 0% |
| 202229 | 2 | 0% | 16% | 84% | 0% | 0% | 0% | 0% | 0% | 0% | 0% | 0% | 0% | 0% | 0% | 0% | 0% | 0% | 0% | 0% |
| 202230 | 2 | 0% | 22% | 78% | 0% | 0% | 0% | 0% | 0% | 0% | 0% | 0% | 0% | 0% | 0% | 0% | 0% | 0% | 0% | 0% |
| 202231 | 2 | 0% | 1% | 99% | 0% | 0% | 0% | 0% | 0% | 0% | 0% | 0% | 0% | 0% | 0% | 0% | 0% | 0% | 0% | 0% |
| 202232 | 0 | - | - | - | - | - | - | - | - | - | - | - | - | - | - | - | - | - | - | - |
| 202233 | 0 | - | - | - | - | - | - | - | - | - | - | - | - | - | - | - | - | - | - | - |
| 202234 | 2 | 0% | 0% | 100% | 0% | 0% | 0% | 0% | 0% | 0% | 0% | 0% | 0% | 0% | 0% | 0% | 0% | 0% | 0% | 0% |
| 202235 | 2 | 0% | 0% | 100% | 0% | 0% | 0% | 0% | 0% | 0% | 0% | 0% | 0% | 0% | 0% | 0% | 0% | 0% | 0% | 0% |
| 202236 | 2 | 0% | 0% | 100% | 0% | 0% | 0% | 0% | 0% | 0% | 0% | 0% | 0% | 0% | 0% | 0% | 0% | 0% | 0% | 0% |
| 202237 | 2 | 0% | 0% | 95% | 0% | 0% | 0% | 0% | 0% | 0% | 0% | 0% | 0% | 0% | 0% | 0% | 0% | 0% | 0% | 5% |
| 202238 | 2 | 0% | 0% | 97% | 0% | 0% | 0% | 0% | 0% | 0% | 0% | 0% | 0% | 0% | 0% | 0% | 0% | 0% | 0% | 3% |
| 202239 | 2 | 0% | 0% | 100% | 0% | 0% | 0% | 0% | 0% | 0% | 0% | 0% | 0% | 0% | 0% | 0% | 0% | 0% | 0% | 0% |
| 202240 | 1 | 0% | 0% | 100% | 0% | 0% | 0% | 0% | 0% | 0% | 0% | 0% | 0% | 0% | 0% | 0% | 0% | 0% | 0% | 0% |
| 202241 | 2 | 0% | 0% | 100% | 0% | 0% | 0% | 0% | 0% | 0% | 0% | 0% | 0% | 0% | 0% | 0% | 0% | 0% | 0% | 0% |
| 202242 | 2 | 0% | 0% | 100% | 0% | 0% | 0% | 0% | 0% | 0% | 0% | 0% | 0% | 0% | 0% | 0% | 0% | 0% | 0% | 0% |
| 202243 | 2 | 0% | 0% | 98% | 0% | 0% | 0% | 0% | 0% | 0% | 0% | 0% | 0% | 0% | 0% | 0% | 0% | 0% | 0% | 2% |
| 202244 | 2 | 0% | 0% | 100% | 0% | 0% | 0% | 0% | 0% | 0% | 0% | 0% | 0% | 0% | 0% | 0% | 0% | 0% | 0% | 0% |
| 202245 | 2 | 0% | 0% | 82% | 0% | 11% | 0% | 7% | 0% | 0% | 0% | 0% | 0% | 0% | 0% | 0% | 0% | 0% | 0% | 0% |
| 202246 | 2 | 0% | 0% | 86% | 0% | 7% | 0% | 6% | 0% | 0% | 0% | 0% | 0% | 0% | 0% | 0% | 0% | 0% | 0% | 0% |
| 202247 | 2 | 0% | 1% | 82% | 0% | 7% | 0% | 10% | 0% | 0% | 0% | 0% | 0% | 0% | 0% | 0% | 0% | 0% | 0% | 0% |
| 202248 | 2 | 0% | 0% | 86% | 0% | 5% | 0% | 8% | 0% | 0% | 0% | 0% | 0% | 0% | 0% | 0% | 0% | 0% | 0% | 1% |
| 202249 | 2 | 0% | 0% | 88% | 0% | 5% | 0% | 7% | 0% | 0% | 0% | 0% | 0% | 0% | 0% | 0% | 0% | 0% | 0% | 0% |
| 202250 | 2 | 0% | 0% | 73% | 0% | 21% | 0% | 7% | 0% | 0% | 0% | 0% | 0% | 0% | 0% | 0% | 0% | 0% | 0% | 0% |
| 202251 | 2 | 0% | 0% | 91% | 0% | 9% | 0% | 0% | 0% | 0% | 0% | 0% | 0% | 0% | 0% | 0% | 0% | 0% | 0% | 0% |
| 202252 | 2 | 0% | 1% | 87% | 0% | 3% | 0% | 9% | 0% | 0% | 0% | 0% | 0% | 0% | 0% | 0% | 0% | 0% | 0% | 0% |

| | | | | | | | | | | | | | | | | | | | | |
|---|---|---|---|---|---|---|---|---|---|---|---|---|---|---|---|---|---|---|---|---|
| 202301 | 0 | - | - | - | - | - | - | - | - | - | - | - | - | - | - | - | - | - | - | - |
| 202302 | 2 | 0% | 0% | 91% | 0% | 8% | 0% | 0% | 0% | 0% | 0% | 0% | 0% | 0% | 0% | 0% | 0% | 0% | 0% | 1% |
| 202303 | 2 | 0% | 0% | 99% | 0% | 1% | 0% | 0% | 0% | 0% | 0% | 0% | 0% | 0% | 0% | 0% | 0% | 0% | 0% | 0% |
| 202304 | 3 | 0% | 0% | 91% | 0% | 2% | 0% | 7% | 0% | 0% | 0% | 0% | 0% | 0% | 0% | 0% | 0% | 0% | 0% | 1% |
| 202305 | 1 | 0% | 20% | 59% | 0% | 17% | 0% | 4% | 0% | 0% | 0% | 0% | 0% | 0% | 0% | 0% | 0% | 0% | 0% | 0% |
| 202306 | 3 | 0% | 5% | 61% | 0% | 17% | 6% | 5% | 6% | 0% | 0% | 0% | 0% | 0% | 0% | 0% | 0% | 0% | 0% | 0% |
| 202307 | 2 | 0% | 0% | 18% | 0% | 14% | 65% | 3% | 0% | 1% | 0% | 0% | 0% | 0% | 0% | 0% | 0% | 0% | 0% | 0% |
| 202308 | 1 | 0% | 0% | 0% | 0% | 0% | 82% | 18% | 0% | 0% | 0% | 0% | 0% | 0% | 0% | 0% | 0% | 0% | 0% | 0% |
| 202309 | 2 | 0% | 0% | 7% | 0% | 55% | 7% | 0% | 6% | 5% | 0% | 0% | 0% | 0% | 0% | 0% | 16% | 0% | 0% | 3% |
| 202310 | 1 | 0% | 0% | 0% | 0% | 0% | 0% | 0% | 0% | 100% | 0% | 0% | 0% | 0% | 0% | 0% | 0% | 0% | 0% | 0% |
| 202311 | 1 | 0% | 0% | 8% | 0% | 13% | 15% | 0% | 0% | 64% | 0% | 0% | 0% | 0% | 0% | 0% | 0% | 0% | 0% | 0% |
| 202312 | 1 | 0% | 0% | 22% | 0% | 78% | 0% | 0% | 0% | 0% | 0% | 0% | 0% | 0% | 0% | 0% | 0% | 0% | 0% | 0% |
| 202313 | 1 | 0% | 0% | 14% | 0% | 6% | 7% | 0% | 0% | 73% | 0% | 0% | 0% | 0% | 0% | 0% | 0% | 0% | 0% | 0% |
| 202314 | 1 | 0% | 0% | 0% | 0% | 0% | 17% | 0% | 0% | 83% | 0% | 0% | 0% | 0% | 0% | 0% | 0% | 0% | 0% | 0% |
| 202315 | 0 | - | - | - | - | - | - | - | - | - | - | - | - | - | - | - | - | - | - | - |
| 202316 | 0 | - | - | - | - | - | - | - | - | - | - | - | - | - | - | - | - | - | - | - |
| 202317 | 0 | - | - | - | - | - | - | - | - | - | - | - | - | - | - | - | - | - | - | - |
| 202318 | 0 | - | - | - | - | - | - | - | - | - | - | - | - | - | - | - | - | - | - | - |
| 202319 | 0 | - | - | - | - | - | - | - | - | - | - | - | - | - | - | - | - | - | - | - |
| 202320 | 0 | - | - | - | - | - | - | - | - | - | - | - | - | - | - | - | - | - | - | - |
| 202321 | 2 | 0% | 0% | 0% | 0% | 0% | 2% | 0% | 7% | 64% | 0% | 0% | 24% | 0% | 0% | 0% | 0% | 0% | 0% | 3% |
| 202322 | 2 | 0% | 0% | 0% | 0% | 6% | 0% | 0% | 1% | 71% | 2% | 0% | 20% | 0% | 0% | 0% | 0% | 0% | 0% | 0% |
| 202323 | 2 | 0% | 0% | 0% | 0% | 7% | 0% | 0% | 4% | 62% | 0% | 0% | 22% | 0% | 0% | 3% | 0% | 0% | 0% | 1% |
| 202324 | 2 | 0% | 0% | 0% | 0% | 28% | 0% | 0% | 0% | 32% | 0% | 0% | 38% | 0% | 0% | 3% | 0% | 0% | 0% | 0% |
| 202325 | 2 | 0% | 0% | 0% | 0% | 0% | 0% | 0% | 8% | 36% | 0% | 0% | 21% | 0% | 0% | 26% | 0% | 0% | 0% | 9% |
| 202326 | 3 | 0% | 0% | 0% | 0% | 10% | 0% | 0% | 0% | 47% | 0% | 0% | 14% | 0% | 0% | 25% | 0% | 0% | 0% | 3% |

| | | | | | | | | | | | | | | | | | | | | |
|---|---|---|---|---|---|---|---|---|---|---|---|---|---|---|---|---|---|---|---|---|
| 202327 | 1 | 0% | 0% | 0% | 0% | 15% | 0% | 0% | 0% | 10% | 0% | 0% | 69% | 0% | 0% | 4% | 0% | 0% | 0% | 3% |
| 202328 | 2 | 0% | 0% | 0% | 0% | 14% | 0% | 0% | 0% | 40% | 0% | 0% | 40% | 0% | 0% | 6% | 0% | 0% | 0% | 0% |
| 202329 | 2 | 0% | 0% | 0% | 0% | 2% | 0% | 0% | 0% | 57% | 0% | 0% | 24% | 0% | 0% | 17% | 0% | 0% | 0% | 0% |
| 202330 | 2 | 0% | 0% | 0% | 0% | 2% | 0% | 0% | 0% | 87% | 0% | 0% | 11% | 0% | 0% | 0% | 0% | 0% | 0% | 1% |
| 202331 | 2 | 0% | 0% | 0% | 0% | 3% | 0% | 0% | 0% | 37% | 0% | 0% | 27% | 0% | 0% | 22% | 3% | 0% | 0% | 6% |
| 202332 | 2 | 0% | 0% | 0% | 0% | 0% | 0% | 0% | 0% | 54% | 0% | 0% | 27% | 0% | 0% | 13% | 0% | 0% | 0% | 6% |
| 202333 | 0 | - | - | - | - | - | - | - | - | - | - | - | - | - | - | - | - | - | - | - |
| 202334 | 2 | 0% | 0% | 0% | 0% | 7% | 0% | 0% | 0% | 3% | 0% | 0% | 43% | 0% | 0% | 42% | 0% | 0% | 0% | 4% |
| 202335 | 2 | 0% | 0% | 0% | 0% | 2% | 0% | 0% | 0% | 1% | 1% | 0% | 27% | 29% | 6% | 30% | 0% | 1% | 0% | 4% |
| 202336 | 2 | 0% | 0% | 1% | 0% | 1% | 0% | 0% | 0% | 0% | 0% | 0% | 20% | 25% | 13% | 37% | 2% | 0% | 0% | 0% |
| 202337 | 2 | 0% | 0% | 0% | 0% | 0% | 0% | 0% | 0% | 1% | 15% | 0% | 8% | 33% | 4% | 39% | 0% | 0% | 0% | 0% |
| 202338 | 2 | 0% | 0% | 0% | 0% | 3% | 0% | 0% | 0% | 0% | 0% | 1% | 16% | 18% | 16% | 37% | 1% | 7% | 0% | 1% |
| 202339 | 2 | 0% | 0% | 0% | 0% | 0% | 0% | 0% | 0% | 2% | 4% | 0% | 14% | 21% | 18% | 41% | 0% | 0% | 0% | 0% |
| 202340 | 2 | 0% | 0% | 0% | 0% | 0% | 0% | 0% | 0% | 1% | 0% | 2% | 12% | 10% | 20% | 47% | 0% | 0% | 0% | 9% |
| 202341 | 1 | 0% | 0% | 0% | 0% | 0% | 0% | 0% | 0% | 0% | 0% | 4% | 3% | 8% | 34% | 12% | 0% | 40% | 0% | 0% |
| 202342 | 2 | 0% | 0% | 0% | 0% | 0% | 0% | 0% | 0% | 19% | 0% | 0% | 22% | 8% | 21% | 23% | 0% | 2% | 5% | 0% |
| 202343 | 2 | 0% | 0% | 0% | 0% | 0% | 0% | 0% | 0% | 0% | 0% | 0% | 16% | 8% | 28% | 19% | 0% | 28% | 0% | 0% |
| 202344 | 1 | 0% | 0% | 0% | 0% | 0% | 0% | 0% | 0% | 0% | 2% | 2% | 12% | 1% | 54% | 8% | 0% | 15% | 6% | 1% |
| 202345 | 1 | 0% | 0% | 0% | 0% | 0% | 0% | 0% | 0% | 0% | 0% | 4% | 3% | 0% | 47% | 16% | 0% | 21% | 10% | 0% |
| 202346 | 1 | 0% | 0% | 0% | 0% | 0% | 0% | 0% | 0% | 0% | 0% | 7% | 12% | 8% | 53% | 5% | 0% | 16% | 0% | 0% |
| 202347 | 1 | 0% | 0% | 0% | 0% | 0% | 0% | 0% | 0% | 1% | 0% | 6% | 0% | 2% | 7% | 5% | 0% | 21% | 59% | 0% |
| 202348 | 1 | 0% | 0% | 0% | 0% | 7% | 0% | 0% | 0% | 0% | 2% | 50% | 10% | 4% | 10% | 3% | 0% | 8% | 6% | 0% |
| 202349 | 1 | 0% | 0% | 0% | 0% | 0% | 0% | 0% | 0% | 10% | 0% | 0% | 4% | 0% | 61% | 8% | 0% | 17% | 0% | 0% |
| 202350 | 2 | 0% | 0% | 0% | 0% | 0% | 0% | 0% | 0% | 1% | 4% | 1% | 1% | 1% | 26% | 1% | 0% | 16% | 50% | 0% |
| 202351 | 2 | 0% | 0% | 0% | 0% | 0% | 0% | 0% | 0% | 9% | 1% | 3% | 0% | 0% | 15% | 1% | 0% | 8% | 63% | 0% |
| 202352 | 0 | - | - | - | - | - | - | - | - | - | - | - | - | - | - | - | - | - | - | - |

| | | | | | | | | | | | | | | | | | | | | |
|---|---|---|---|---|---|---|---|---|---|---|---|---|---|---|---|---|---|---|---|---|
| 202401 | 0 | - | - | - | - | - | - | - | - | - | - | - | - | - | - | - | - | - | - | - |
| 202402 | 2 | 0% | 0% | 0% | 0% | 0% | 0% | 0% | 0% | 1% | 1% | 25% | 0% | 0% | 14% | 0% | 0% | 11% | 48% | 0% |
| 202403 | 2 | 0% | 0% | 0% | 0% | 0% | 0% | 0% | 0% | 2% | 0% | 2% | 0% | 0% | 12% | 0% | 0% | 14% | 69% | 0% |
| 202404 | 2 | 0% | 0% | 0% | 0% | 0% | 0% | 0% | 0% | 0% | 0% | 1% | 0% | 0% | 8% | 5% | 0% | 19% | 66% | 0% |
| 202405 | 2 | 0% | 0% | 0% | 0% | 0% | 0% | 0% | 0% | 1% | 0% | 0% | 0% | 0% | 11% | 0% | 0% | 11% | 77% | 0% |
| 202406 | 2 | 0% | 0% | 0% | 0% | 0% | 0% | 0% | 0% | 0% | 0% | 0% | 0% | 0% | 2% | 0% | 0% | 42% | 56% | 0% |
| 202407 | 2 | 0% | 0% | 0% | 0% | 0% | 0% | 0% | 0% | 0% | 0% | 0% | 0% | 0% | 2% | 0% | 0% | 26% | 72% | 0% |
| 202408 | 2 | 0% | 0% | 0% | 0% | 0% | 0% | 0% | 0% | 1% | 0% | 0% | 2% | 0% | 2% | 0% | 0% | 18% | 78% | 0% |
| 202409 | 2 | 0% | 0% | 0% | 0% | 0% | 0% | 0% | 0% | 2% | 0% | 0% | 0% | 0% | 4% | 0% | 0% | 43% | 52% | 0% |
| 202410 | 2 | 0% | 0% | 0% | 0% | 0% | 0% | 0% | 0% | 0% | 0% | 0% | 0% | 0% | 3% | 0% | 0% | 24% | 73% | 0% |
| 202411 | 2 | 0% | 0% | 0% | 0% | 0% | 0% | 0% | 0% | 0% | 0% | 0% | 0% | 0% | 0% | 0% | 0% | 36% | 64% | 0% |
| 202412 | 2 | 0% | 0% | 0% | 0% | 0% | 0% | 0% | 0% | 0% | 0% | 0% | 0% | 0% | 1% | 0% | 0% | 64% | 35% | 0% |
| 202413 | 2 | 0% | 0% | 0% | 0% | 0% | 0% | 0% | 0% | 0% | 0% | 0% | 0% | 0% | 1% | 0% | 0% | 55% | 45% | 0% |

N: number of samples.

Table S4. Unstandardized partial regression coefficients for the incidence of hospitalized cases and $R^2$ for incidence and predicted prevalence.

| | Length of hospital stay (#) | | | | | | | | | | | |
|---|---|---|---|---|---|---|---|---|---|---|---|---|
| | 4 & 3 weeks (main analysis) | | 1 week | | 2 weeks | | 3 weeks | | 4 weeks | | 5 weeks | |
| | B (95% CI) | *P* | B (95% CI) | *P* | B (95% CI) | *P* | B (95% CI) | *P* | B (95% CI) | *P* | B (95% CI) | *P* |
| Constant | −0.067 (−0.428–0.294) | 0.713 | 0.936 (0.749–1.123) | <0.001 | 0.571 (0.390–0.752) | <0.001 | 0.312 (0.067–0.557) | 0.013 | −0.033 (−0.554–0.488) | 0.899 | −0.225 (−0.850–0.400) | 0.477 |
| $\log_{10}$ SARS-CoV-2 RNA (same-week) | 0.278 (0.122–0.434) | <0.001 | 0.178 (0.097–0.259) | <0.001 | 0.221 (0.142–0.299) | <0.001 | 0.263 (0.157–0.369) | <0.001 | 0.354 (0.128–0.580) | 0.002 | 0.352 (0.081–0.623) | 0.011 |
| $\log_{10}$ SARS-CoV-2 RNA (one-week prior) | 0.406 (0.255–0.558) | <0.001 | 0.411 (0.332–0.490) | <0.001 | 0.386 (0.310–0.462) | <0.001 | 0.364 (0.261–0.467) | <0.001 | 0.320 (0.101–0.539) | 0.005 | 0.342 (0.079–0.605) | 0.011 |
| Wave A (ref: Wave C) | −0.018 (−0.172–0.137) | 0.822 | −0.058 (−0.138–0.022) | 0.154 | −0.053 (−0.130–0.025) | 0.179 | −0.054 (−0.159–0.051) | 0.313 | −0.023 (−0.246–0.201) | 0.841 | −0.078 (−0.346–0.190) | 0.566 |
| Wave B (ref: Wave C) | −0.006 (−0.175–0.164) | 0.949 | −0.084 (−0.172–0.004) | 0.062 | −0.069 (−0.154–0.016) | 0.112 | −0.052 (−0.168–0.063) | 0.369 | −0.001 (−0.247–0.244) | 0.991 | −0.017 (−0.312–0.277) | 0.909 |
| Wave D (ref: Wave C) | −0.015 (−0.166–0.136) | 0.843 | −0.187 (−0.266–−0.109) | <0.001 | −0.188 (−0.264–−0.112) | <0.001 | −0.190 (−0.292–−0.087) | <0.001 | −0.204 (−0.422–0.014) | 0.066 | −0.186 (−0.448–0.075) | 0.161 |
| Wave E (ref: Wave C) | −0.044 (−0.214–0.127) | 0.611 | −0.220 (−0.308–−0.131) | <0.001 | −0.212 (−0.297–−0.126) | <0.001 | −0.218 (−0.334–−0.102) | <0.001 | −0.232 (−0.478–0.015) | 0.065 | −0.233 (−0.529–0.063) | 0.121 |
| $R^2$ for incidence of cases | 0.6125 | | 0.8075 | | 0.8244 | | 0.7310 | | 0.4110 | | 0.3402 | |
| $R^2$ for prevalence | 0.9666 | | 0.8075 | | 0.9526 | | 0.9667 | | 0.9668 | | 0.9641 | |

B: unstandardized partial regression coefficient. CI: confidence interval.
(#) Under main analysis, the length of hospital stay was set to 4 and 3 weeks for the pre- and post-reclassification periods, respectively.
For sensitivity analyses, the length of hospital stay was set to 1–5 weeks for both the pre- and post-reclassification periods.

Table S5. Unstandardized partial regression coefficients for incidence of severe cases and $R^2$ for incidence and predicted prevalence.

| | Length of hospital stay (#) | | | | | | | | | | | |
|---|---|---|---|---|---|---|---|---|---|---|---|---|
| | 5 & 4 weeks (main analysis) | | 2 weeks | | 3 weeks | | 4 weeks | | 5 weeks | | 6 weeks | |
| | B (95% CI) | *P* | B (95% CI) | *P* | B (95% CI) | *P* | B (95% CI) | *P* | B (95% CI) | *P* | B (95% CI) | *P* |
| Constant | −1.110 (−1.707–−0.512) | <0.001 | −1.381 (−1.827–−0.935) | <0.001 | −1.179 (−1.703–−0.656) | <0.001 | −1.113 (−1.695–−0.532) | <0.001 | −0.992 (−1.586–−0.398) | 0.001 | −0.949 (−1.702–−0.196) | 0.014 |
| $\log_{10}$ SARS-CoV-2 RNA (same-week) | 0.135 (−0.124–0.394) | 0.304 | 0.162 (−0.031–0.355) | 0.099 | 0.111 (−0.115–0.338) | 0.332 | 0.127 (−0.125–0.378) | 0.321 | 0.138 (−0.119–0.395) | 0.290 | 0.182 (−0.144–0.508) | 0.272 |
| $\log_{10}$ SARS-CoV-2 RNA (one-week prior) | 0.373 (0.121–0.624) | 0.004 | 0.542 (0.355–0.730) | <0.001 | 0.479 (0.259–0.699) | <0.001 | 0.403 (0.158–0.647) | 0.001 | 0.335 (0.085–0.585) | 0.009 | 0.247 (−0.070–0.564) | 0.125 |
| Wave A (ref: Wave C) | 0.063 (−0.193–0.319) | 0.627 | 0.148 (−0.043–0.339) | 0.128 | 0.104 (−0.120–0.328) | 0.358 | 0.098 (−0.151–0.347) | 0.436 | 0.061 (−0.193–0.315) | 0.636 | 0.000 (−0.323–0.322) | 0.998 |
| Wave B (ref: Wave C) | −0.045 (−0.327–0.236) | 0.749 | 0.023 (−0.187–0.233) | 0.831 | 0.032 (−0.214–0.279) | 0.795 | 0.012 (−0.262–0.285) | 0.933 | −0.038 (−0.318–0.242) | 0.789 | −0.054 (−0.409–0.301) | 0.764 |
| Wave D (ref: Wave C) | −0.151 (−0.401–0.100) | 0.236 | −0.163 (−0.350–0.023) | 0.086 | −0.223 (−0.442–−0.003) | 0.047 | −0.216 (−0.459–0.027) | 0.081 | −0.233 (−0.482–0.015) | 0.066 | −0.312 (−0.627–0.004) | 0.053 |
| Wave E (ref: Wave C) | −0.356 (−0.638–−0.073) | 0.014 | −0.543 (−0.754–−0.332) | <0.001 | −0.467 (−0.714–−0.219) | <0.001 | −0.427 (−0.701–−0.152) | 0.003 | −0.381 (−0.662–−0.100) | 0.008 | −0.389 (−0.745–−0.033) | 0.032 |
| $R^2$ for incidence of cases | 0.2506 | | 0.5589 | | 0.4007 | | 0.3010 | | 0.2399 | | 0.1452 | |
| $R^2$ for prevalence | 0.8199 | | 0.8304 | | 0.8386 | | 0.8213 | | 0.8170 | | 0.7984 | |

B: unstandardized partial regression coefficient. CI: confidence interval.
(#) Under main analysis, the length of hospital stay was set to 5 and 4 weeks for the pre- and post-reclassification periods, respectively.
For sensitivity analyses, the length of hospital stay was set to 2–6 weeks for both the pre- and post-reclassification periods.

Table S6. Unstandardized partial regression coefficients for the incidence of confirmed, hospitalized, and severe cases by using PMMoV corrections.

| | Incidence of confirmed cases (Model 3) | | Incidence of hospitalized cases (Model 2) | | Incidence of severe cases (Model 2) | |
|---|---|---|---|---|---|---|
| | B (95% CI) | *P* | B (95% CI) | *P* | B (95% CI) | *P* |
| Constant | 6.918 (6.719–7.117) | <0.001 | 4.488 (4.122–4.854) | <0.001 | 2.293 (1.672–2.913) | <0.001 |
| $log_{10}$ SARS-CoV-2 RNA/PMMoV RNA (same-week) | 0.423 (0.333–0.514) | <0.001 | 0.289 (0.123–0.455) | <0.001 | 0.117 (−0.165–0.399) | 0.412 |
| $log_{10}$ SARS-CoV-2 RNA/PMMoV RNA (one-week prior) | 0.296 (0.188–0.404) | <0.001 | 0.415 (0.253–0.577) | <0.001 | 0.412 (0.138–0.686) | 0.004 |
| $log_{10}$ SARS-CoV-2 RNA/PMMoV RNA (two-week prior) | 0.107 (0.019–0.195) | 0.018 | - | - | - | - |
| Wave A (ref: Wave C) | 0.212 (0.132–0.293) | <0.001 | 0.049 (−0.100–0.199) | 0.514 | 0.114 (−0.139–0.367) | 0.373 |
| Wave B (ref: Wave C) | 0.214 (0.126–0.302) | <0.001 | 0.002 (−0.162–0.165) | 0.984 | −0.039 (−0.317–0.238) | 0.780 |
| Wave D (ref: Wave C) | −3.567 (−3.644–−3.489) | <0.001 | 0.007 (−0.138–0.152) | 0.924 | −0.134 (−0.380–0.112) | 0.283 |
| Wave E (ref: Wave C) | −3.836 (−3.925–−3.747) | <0.001 | −0.076 (−0.242–0.089) | 0.363 | −0.380 (−0.661–−0.100) | 0.008 |

B: unstandardized partial regression coefficient. CI: confidence interval.

Table S7. Comparison of multiple regression models using variant proportion and wave dummy variables.

| | Incidence of confirmed cases (Model 3) | | | Incidence of hospitalized cases (Model 2) | | | Incidence of severe cases (Model 2) | | |
|---|---|---|---|---|---|---|---|---|---|
| | Variant (multi-week average) | Variant (strong association week) | Wave dummy | Variant (multi-week average) | Variant (strong association week) | Wave dummy | Variant (multi-week average) | Variant (strong association week) | Wave dummy |
| $R^2$ | 0.9968 | 0.9961 | 0.9940 | 0.6736 | 0.6855 | 0.6250 | 0.3265 | 0.3561 | 0.2246 |
| AIC | −145.48 | −121.47 | −103.71 | 47.93 | 43.76 | 35.63 | 168.29 | 163.20 | 156.21 |
| BIC | −80.03 | −56.01 | −79.16 | 107.93 | 103.76 | 57.45 | 228.29 | 223.20 | 178.03 |

AIC: Akaike information criterion. BIC: Bayesian information criterion.

$R^2$, AIC, and BIC values from the regression analysis using wave dummies differed from those in Table 1 due to differences in the analysis periods.

Table S8. Unstandardized partial regression coefficients for the incidence of confirmed, hospitalized, and severe cases based on regression models fitted to Waves A–D.

| | Incidence of confirmed cases (Model 3) | | Incidence of hospitalized cases (Model 2) | | Incidence of severe cases (Model 2) | |
|---|---|---|---|---|---|---|
| | B (95% CI) | *P* | B (95% CI) | *P* | B (95% CI) | *P* |
| Constant | 1.558 (1.345–1.772) | <0.001 | −0.071 (−0.449–0.307) | 0.710 | −1.074 (−1.677–−0.471) | <0.001 |
| $\log_{10}$ SARS-CoV-2 RNA (same-week) | 0.403 (0.308–0.498) | <0.001 | 0.263 (0.095–0.432) | 0.003 | 0.240 (−0.029–0.509) | 0.079 |
| $\log_{10}$ SARS-CoV-2 RNA (one-week prior) | 0.301 (0.192–0.410) | <0.001 | 0.422 (0.258–0.587) | <0.001 | 0.257 (−0.006–0.520) | 0.055 |
| $\log_{10}$ SARS-CoV-2 RNA (two-week prior) | 0.103 (0.011–0.196) | 0.029 | - | - | - | - |
| Wave A (ref: Wave C) | 0.134 (0.046–0.221) | 0.003 | −0.017 (−0.175–0.141) | 0.835 | 0.056 (−0.196–0.308) | 0.660 |
| Wave B (ref: Wave C) | 0.205 (0.109–0.301) | <0.001 | −0.006 (−0.180–0.168) | 0.945 | −0.041 (−0.318–0.237) | 0.771 |
| Wave D (ref: Wave C) | −3.593 (−3.679–−3.508) | <0.001 | −0.015 (−0.170–0.139) | 0.847 | −0.150 (−0.397–0.097) | 0.230 |

B: unstandardized partial regression coefficient. CI: confidence interval.

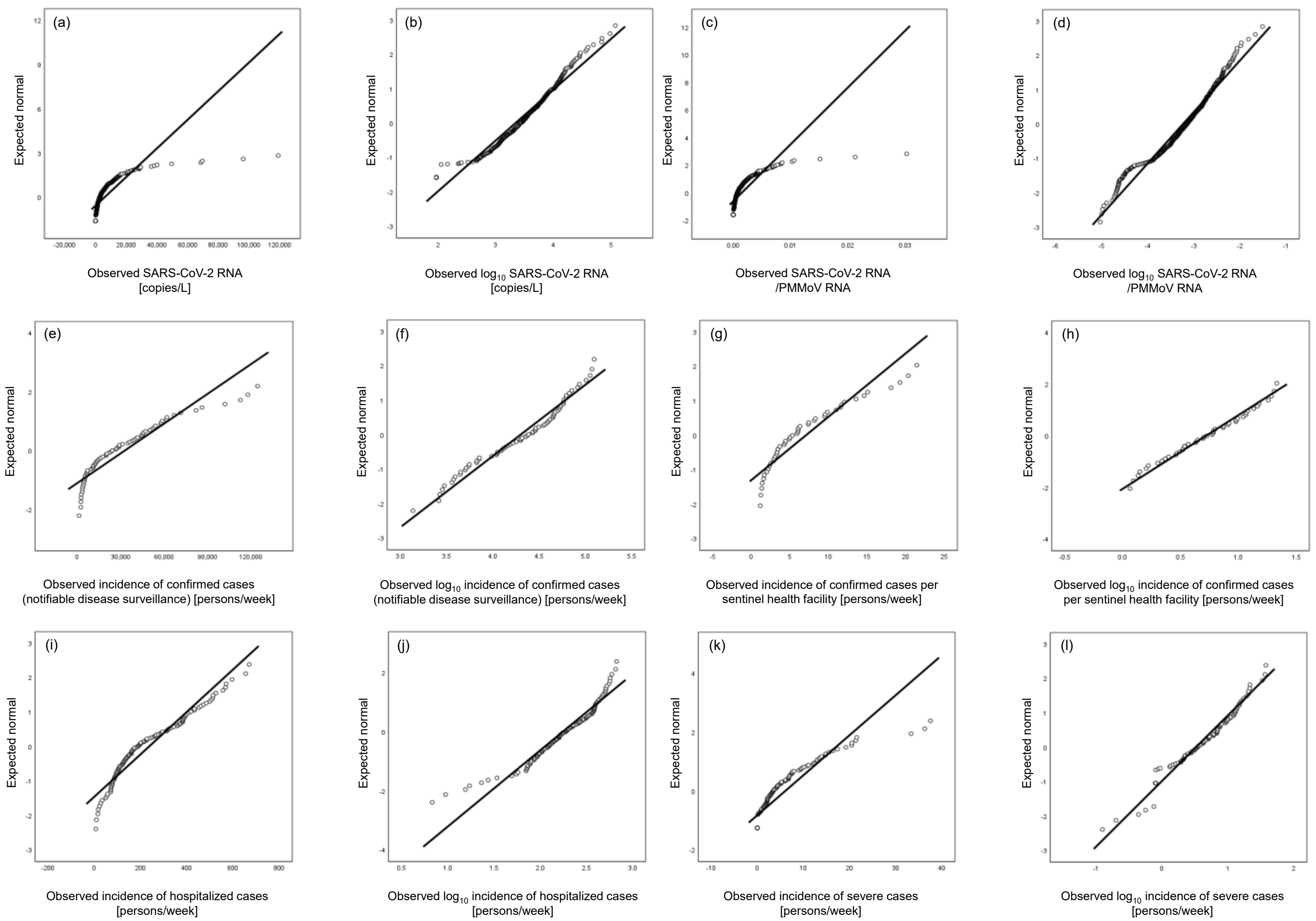


Figure S1. Q–Q plots for SARS-CoV-2 RNA concentrations, SARS-CoV-2 RNA/PMMoV, incidence of confirmed, hospitalized, and severe cases. (a) SARS-CoV-2 RNA concentrations; (b) $\log_{10}$ SARS-CoV-2 RNA concentrations; (c) SARS-CoV-2 RNA/PMMoV RNA ratios; (d) $\log_{10}$ SARS-CoV-2 RNA/PMMoV RNA ratios; (e) incidence of confirmed cases (notifiable disease surveillance); (f) $\log_{10}$ incidence of confirmed cases (notifiable disease surveillance); (g) incidence of confirmed cases per sentinel health facility; (h) $\log_{10}$ incidence of confirmed cases per sentinel health facility; (i) incidence of hospitalized cases; (j) $\log_{10}$ incidence of hospitalized cases; (k) incidence of severe cases; (l) $\log_{10}$ incidence of severe cases.

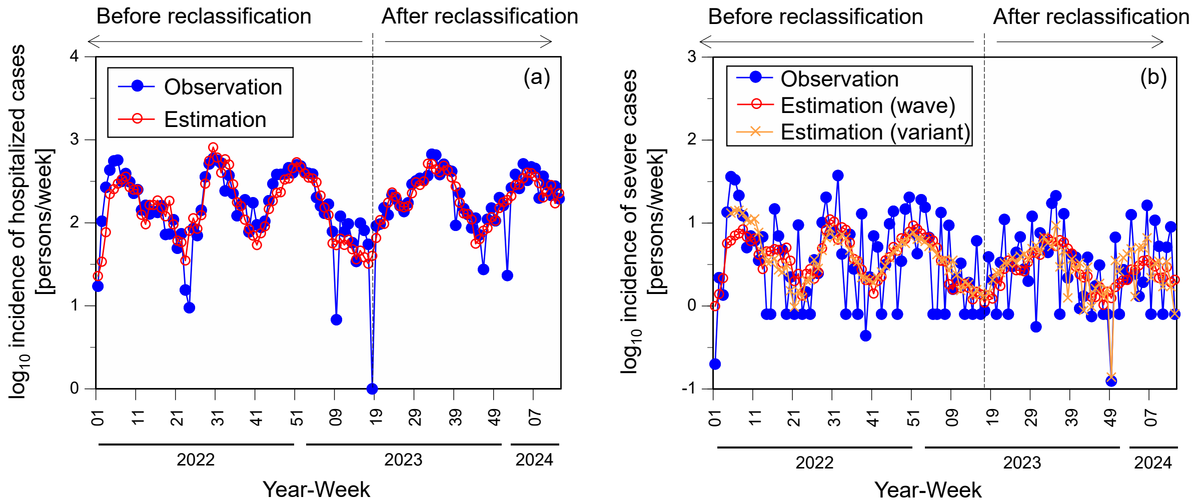


Figure S2. Comparison between observed and estimated values based on regression models using wave dummy and variant proportion variables. (a) Incidence of hospitalized cases (wave dummy: $R^2$ = 0.6125; variant [strong-association week]: 0.6855); (b) incidence of severe cases (wave dummy: $R^2$ = 0.2506; variant [strong-association week]: 0.3561).

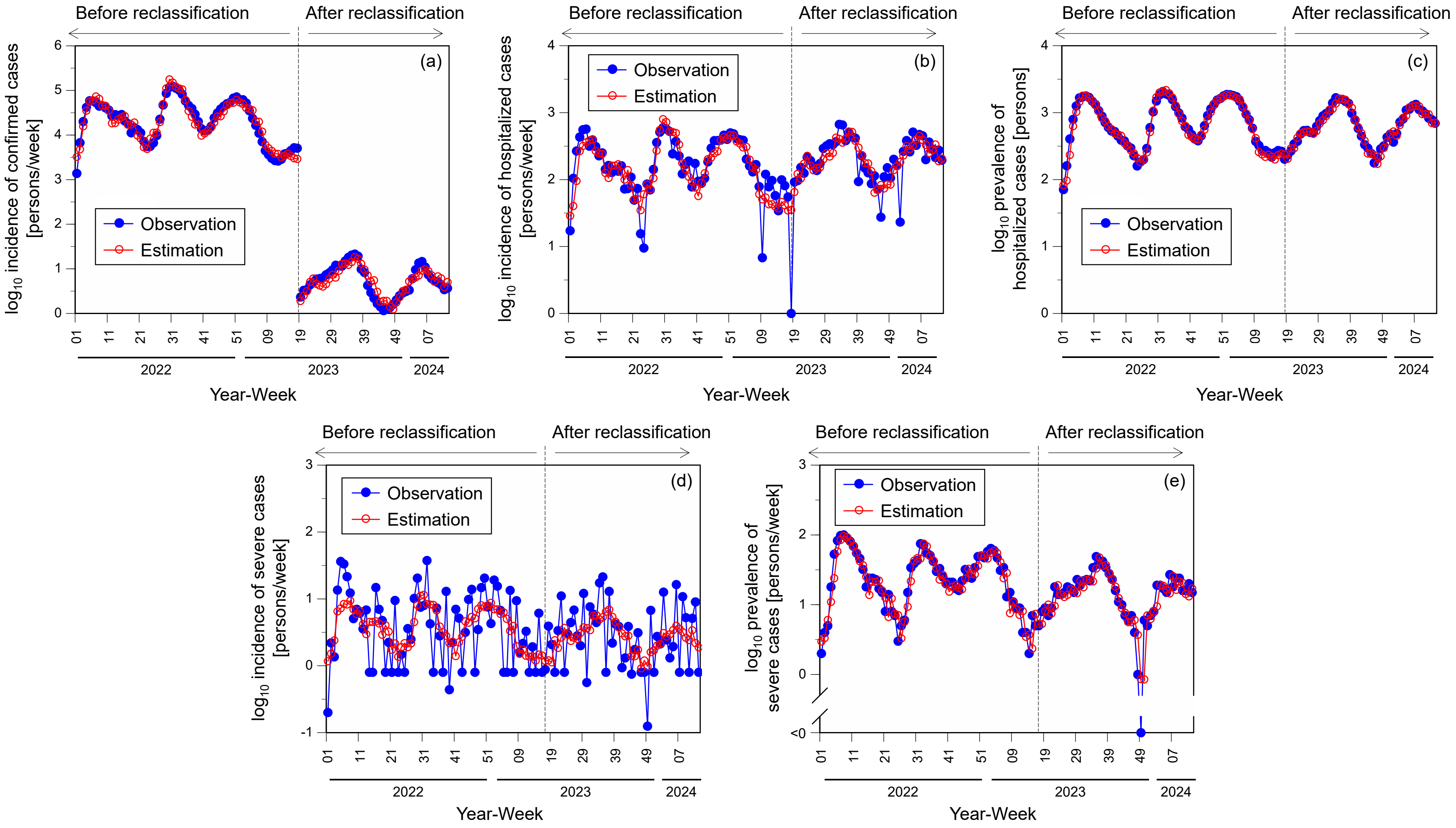


Figure S3. Comparison between observed and estimated values by using PMMoV corrections. (a) Incidence of confirmed cases ($R^2$ = 0.9940 for overall period; 0.9255 for notifiable disease surveillance period; $R^2$ = 0.8133 for sentinel surveillance period); (b) incidence of hospitalized cases ($R^2$ = 0.6391); (c) prevalence of hospitalized cases ($R^2$ = 0.9703); (d) incidence of severe cases ($R^2$ = 0.2698); (e) prevalence of severe cases ($R^2$ = 0.8221 [the calculation excludes data from one week when the number of severe cases was zero]).

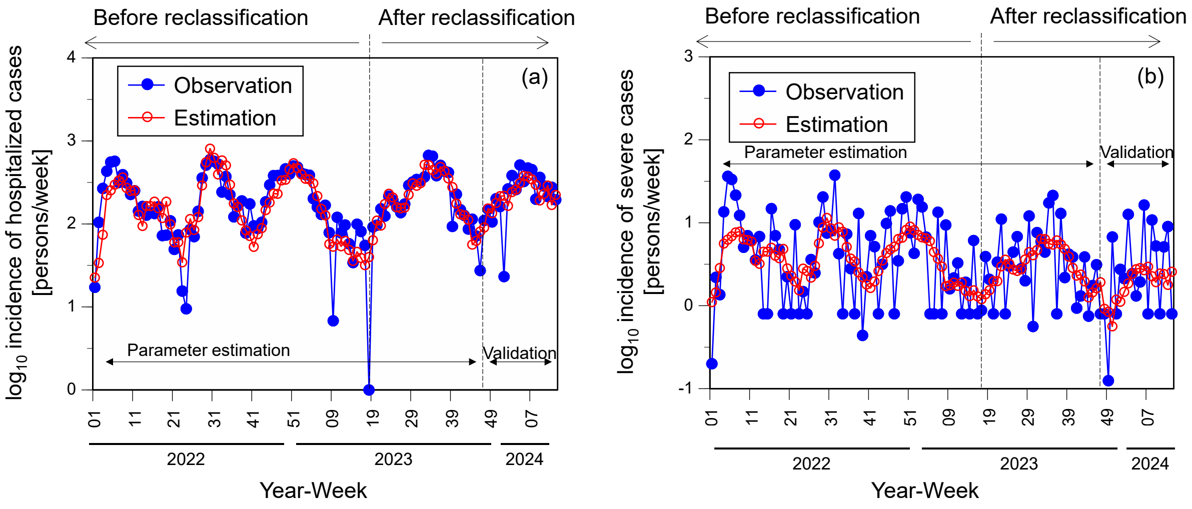


Figure S4. Comparison between observed and estimated values based on regression models fitted to Waves A–D with calibration using data from the validation period (Wave E). (a) Incidence of hospitalized cases (root mean squared error [RMSE] = 0.2764 at parameter estimation period; RMSE = 0.2641 at validation period); (b) incidence of severe cases (RMSE = 0.4414 at parameter estimation period; RMSE = 0.5486 at validation period).
